# Estimating the relative contribution of transmission to bedaquiline resistance burden in tuberculosis. A transmission threshold model

**DOI:** 10.64898/2026.01.11.26343862

**Authors:** Abel Kjaersgaard, Chaelin Kim, Thobani Ntshiqa, Salome Charalambous, C Finn McQuaid, Lara Goscé, Richard G White, Timothy D McHugh, Rein M G J Houben, Gwenan M Knight

## Abstract

Bedaquiline (BDQ) is a cornerstone antibiotic for treating multidrug- or rifampicin-resistant tuberculosis (MDR/RR-TB). Resistance to BDQ (BDQR) can arise through three mechanisms: spontaneous mutation, acquisition during treatment, or transmission of a resistant strain. We developed a transmission threshold model to estimate the contribution of each mechanism to BDQR burden in MDR/RR-TB

We integrated four data sources: (i) novel estimates of spontaneous resistance probability; (ii) updated estimates of acquired resistance during treatment probability; (iii) a systematic review of country-level BDQR prevalence in MDR/RR-TB (BR-MDR/RR-TB); and (iv) WHO MDR/RR-TB notification and treatment data. We estimated the proportion of BR-MDR/RR-TB attributable to transmission, using transmission threshold values defined as the ratio of BDQR observed in prevalence studies to expectations from spontaneous and acquired mechanisms alone. Analyses were conducted across hypothetical BDQR prevalence and treatment coverage scenarios, as well as country-specific data.

We estimated spontaneous resistance among all MDR/RR-TB at 0.073% (95% CrI: 0.062-0.085%) and acquired resistance among MDR/RR-TB patients receiving BDQ at 3.6% (95% CrI: 2.8-4.4%). BDQR prevalence among MDR/RR-TB varied between 0-20% across 18 countries between 2015-2024. Scenario modelling suggested that spontaneous and acquired resistance alone could not account for BDQR prevalence among MDR/RR-TB above 1.9% in settings where ≤50% of MDR/RR-TB patients had received BDQ in the previous year— suggesting possible transmission. In Brazil, China, Mozambique, and South Africa, threshold values and associated uncertainty were consistent with BR-MDR/RR-TB transmission since 2020. Other countries had threshold values consistent with resistance primarily arising spontaneously or through acquisition.

Determining country- and year-specific threshold values may help guide decisions on when to prioritise protecting a new antibiotic by preventing resistance acquisition versus when ongoing transmission requires interventions focused on interrupting transmission.

## Introduction

Bedaquiline (BDQ) is a cornerstone antibiotic in newly recommended all-oral short regimens (e.g., BPaL(M)) for treating multidrug- and rifampicin-resistant tuberculosis (MDR/RR-TB) [1]. In 2023, 400,000 individuals (approximately 3.7% of all TB cases) globally were estimated to have MDR/RR-TB, underscoring the clinical importance of BDQ [2]. As of 2025, increasing levels of BDQ resistance have already been observed in some settings. This highlights the urgent need for interventions that mitigate acquisition and/or transmission of resistance in order to preserve this antibiotic.

Resistance to BDQ in *Mycobacterium tuberculosis (M.tb.)* occurs via three principal mechanisms: spontaneous, acquired, and transmitted. Noticeably, *M.tb.* shows limited evidence of horizontal gene transfer [3,4]. Understanding the relative contribution of each mechanism to BR-MDR/RR-TB burden is critical for informed TB control strategies: e.g., should interventions prioritise safeguarding the new drug against acquired resistance, or should efforts focus on interrupting transmission chains?

Spontaneous resistance occurs when a dividing BDQ-susceptible *M.tb.* strain within a host produces BDQR offspring due to random *de novo* mutations, and the resulting BDQR subpopulation grows sufficiently large to be detectable with drug-susceptibility testing (DST). Acquired resistance occurs in TB patients when the *M.tb.* strain is detected as BDQ-susceptible (BDQS) at treatment baseline, but BDQR at follow-up DST during or after treatment with BDQ. Acquired resistance thus reflects situations where either a small BDQR subpopulation is undetected at baseline, but selected for during treatment, or the entire *M.tb.* population is BDQ-susceptible at baseline, but BDQR mutations arise and are selected for during treatment. Finally, transmitted resistance occurs when individuals are infected with a strain that is already BDQR. As BDQ is principally used for treating MDR/RR-TB, we discuss the above mechanisms as they occur in patients already infected with MDR/RR-TB strains, or, in the case of transmission, of strains that are both MDR/RR and BDQR (BR-MDR/RR-TB).

In this study, we developed a transmission threshold model to evaluate whether spontaneous and acquired resistance alone could account for observed BR-MDR/RR-TB burdens. In doing so, we established transmission threshold values above which transmission may be invoked to explain excess BR-MDR/RR-TB. Rather than relying on whole genome sequencing, or extensive population DST sampling to gauge BDQR transmission, our model leverages open WHO data sources and BDQR prevalence estimates to distinguish between resistance mechanisms, offering critical insights for tailoring context-specific interventions to curb the spread of BDQR.

## Methods

We developed a transmission threshold model to assess whether observed BDQR in MDR/RR-TB patients exceeds levels expected from spontaneous and acquired resistance alone. For each country and year with eligible BDQR prevalence data identified through a systematic review, we calculated threshold values: the ratio of observed to expected BR-MDR/RR-TB.

### Data sources and model parameterisation

The model was parameterised using four data sources: (i) novel estimates of spontaneous resistance probability; (ii) renewed estimates of acquired resistance probability; (iii) a systematic review of country-level BDQR prevalence among notified MDR/RR-TB; and (iv) WHO MDR/RR-TB notification and treatment data (Table 1).

**Table 1:**
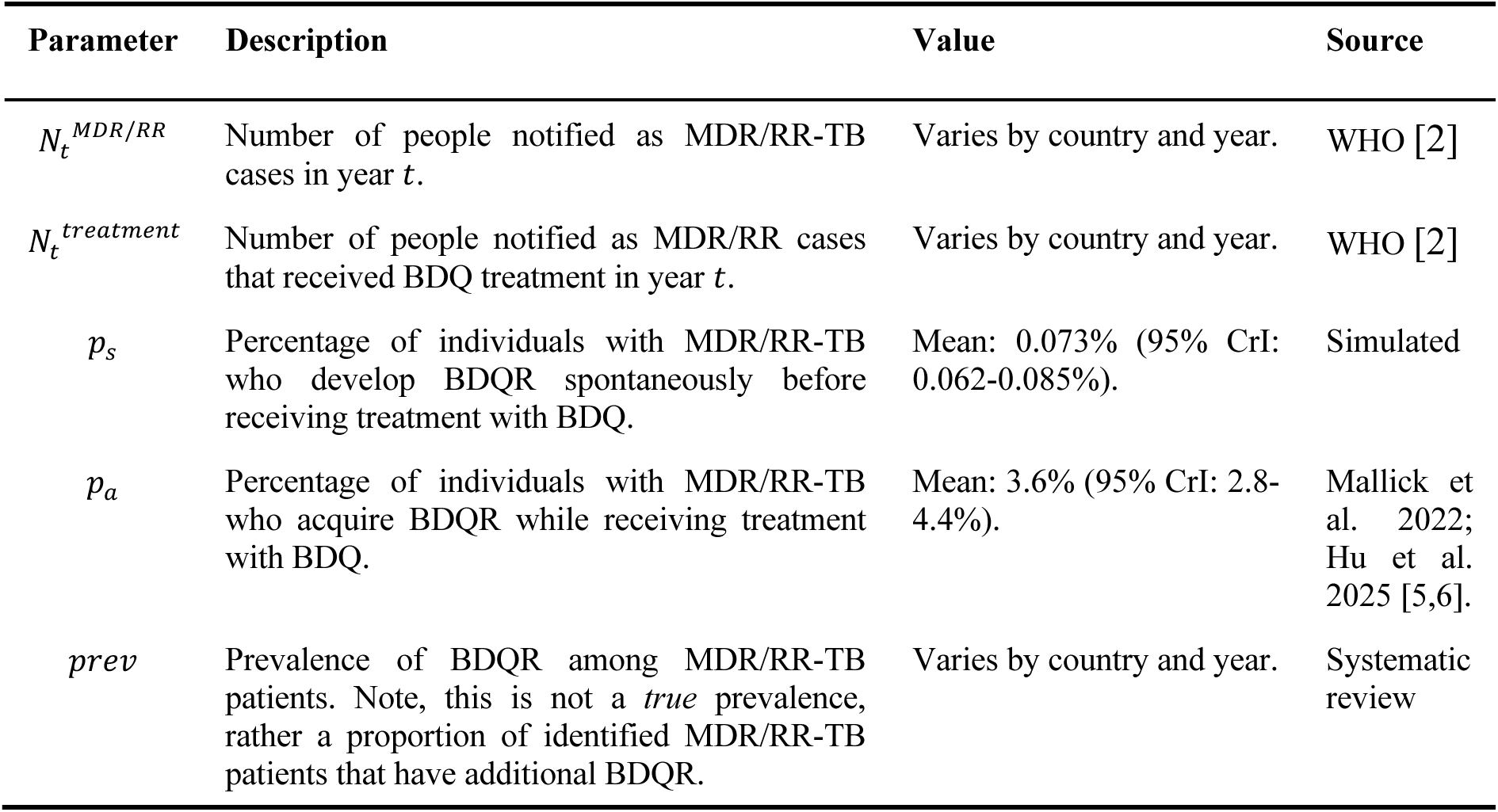
Model parameters are derived from the literature, WHO reports, or simulated in this study. CrI = equal-tailed credible interval.

| Parameter | Description | Value | Source |
| --- | --- | --- | --- |
| $N_t^{MDR/RR}$ | Number of people notified as MDR/RR-TB cases in year $t$. | Varies by country and year. | WHO [2] |
| $N_t^{treatment}$ | Number of people notified as MDR/RR cases that received BDQ treatment in year $t$. | Varies by country and year. | WHO [2] |
| $p_s$ | Percentage of individuals with MDR/RR-TB who develop BDQR spontaneously before receiving treatment with BDQ. | Mean: 0.073% (95% CrI: 0.062-0.085%). | Simulated |
| $p_a$ | Percentage of individuals with MDR/RR-TB who acquire BDQR while receiving treatment with BDQ. | Mean: 3.6% (95% CrI: 2.8-4.4%). | Mallick et al. 2022; Hu et al. 2025 [5,6]. |
| $prev$ | Prevalence of BDQR among MDR/RR-TB patients. Note, this is not a *true* prevalence, rather a proportion of identified MDR/RR-TB patients that have additional BDQR. | Varies by country and year. | Systematic review |

*N*_*t*_^*MDR*/*RR*^, the reported number of individuals with MDR/RR-TB in year *t*, and *N*_*t*_^*treatment*^, the reported number of individuals with MDR/RR-TB receiving BDQ treatment in year *t*, were extracted as point values from WHO data published in 2024 [2]. If *N*_*t*_^*treatment*^ was missing for any country-year where eligible BDQR prevalence data was identified, we imputed missing values conservatively depending on the available data (Supplement S4).

We estimated *p*_*s*_, the probability of spontaneous resistance, using a within-host *M.tb.* simulation, adapted from Colijn et al. (Supplement S1) [7]. We simulated the stochastic growth and emergence of BDQ resistance through *de-novo* mutations in an initially fully BDQ susceptible *M.tb.* population. We assumed the per-division probability of a BDQR-conferring mutation was 5 × 10^−7^—the upper bound of estimates from *in vitro* studies [8]. We assumed that detection and sputum collection would occur when the bacteria population reached 10^10^, with BDQR assumed detectable if ≥ 1% of the bacteria population was resistant—reflecting the limit of detection for phenotypic DST in heteroresistant bacteria populations [9]. We assumed no fitness cost for the BDQR strain [10,11]. We simulated 200,000 independent populations and fitted a Bayesian beta-binomial model to the proportion of simulated populations where resistance was detectable. *p*_*s*_was sampled from the resultant posterior distribution (Supplement S1).

We estimated *p*_*a*_, the probability of acquiring resistance to BDQ during treatment, using pooled data from two systematic reviews. We combine data from 11 studies from a 2022 review by Mallick et al., comprising 1,087 individuals, and a further 5 studies from a 2025 review by Hu et al., comprising 1,056 individuals (Supplement S2) [6,12]. Thus, the final sample contained 2,143 individuals from 16 studies. We extracted the raw incidence data on acquired resistance from each study and fitted a Bayesian hierarchical logistic regression model to account for between-study heterogeneity. We then drew posterior samples from the fitted model and calculated *p*_*a*_ as the study population-weighted mean of the posterior distribution (Supplement S2).

We extracted *prev*, the prevalence of BDQR among incident MDR/RR-TB cases, by conducting a systematic review of published BDQR prevalence studies. We searched PubMed on 26 June 2025, returning 168 articles (including duplicates) (Supplement S3). We did not impose any restrictions on sample size, geographic location, or source of patient cohort (e.g., hospital, national reference laboratory, etc.), or a date range for publications, however, 143/168 (85%) of screened studies were published since 2020. Abstract/title screening was done by two separate reviewers (AK, GK) and excluded 59 references (inclusion/exclusion criteria in Table 2). Full text screening (AK) excluded a further 91, leading to a final sample of 18 studies. We extracted BDQR prevalence among MDR/RR-TB patients from studies where DST results were available at baseline (Table 2). Study summaries are in the supplement (Supplement S3).

**Table 2:** Inclusion/exclusion criteria for systematic review BR-MDR/RR-TB prevalence.

| Stage | Inclusion (must satisfy all) | Exclusion (may not satisfy any) |
| --- | --- | --- |
| Abstract | 1) The study evaluated BDQ or BDQ resistance<br>2) The study recruited a patient cohort (either prospectively or retrospectively) of MDR/RR-TB patients | 1) The study was a literature review, modelling study, case report, *in vitro* study, animal studies, or trial protocols. |
| Full text | 1) TB patients with resistance to at least rifampicin and/or isoniazid<br>2) Patient-level cohort (retrospective or prospective), or baseline of a clinical trial<br>3) Phenotypic or genotypic BDQ resistance testing at baseline (i.e. before BDQ exposure) | 1) Collation of data across studies, without site-level data disaggregation (where possible, data from referenced studies was sourced)<br>2) Non-representative or non-random patient population (e.g., selection on post-treatment outcomes or DST status)<br>3) Did not report BDQ DST data at baseline (or cannot be separated at baseline from other resistance types)<br>4) Did not clearly explain how the comparison (BDQ-S) population was recruited and calculated. |

Some studies spanned multiple years and only reported results in aggregate. In such cases, we assumed MDR/RR-TB and BR-MDR/RR-TB patients to be uniformly distributed over time, and disaggregate these two numbers weighted by the number of months of sampling in each year: *n* × *months of sampling in year*/*total months of sampling in study*). Where this disaggregation was applied, the median number of years smoothed across was two. For each country year, we fitted a Bayesian beta-binomial model to the calculated prevalence and sample size. We then drew posterior samples from the fitted model as *prev* (Table 1).

### Model structure

We assumed that the total number of BR-MDR/RR-TB in a year, *t*, could be disaggregated into three distinct subpopulations: those BDQR due to spontaneous, acquired, and transmitted resistance. Hence:

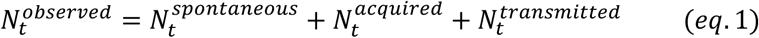

where 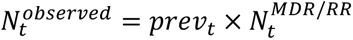, 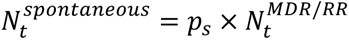 and 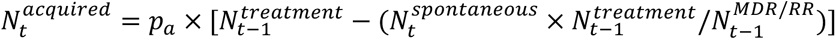 (Figure 1). Note, the population at risk of BDQR due to acquisition in year *t* were all MDR/RR-TB patients receiving treatment in year *t* − 1, less the population that had already developed BDQR spontaneously before starting treatment 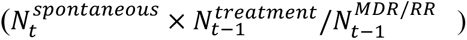. The shortest BDQ treatment course recommended by WHO is currently 6 months [1]. Hence, we assumed that patients treated with BDQ in year *t* − 1 would be detectable as an acquired resistant case in year *t*.

**Figure 1:**
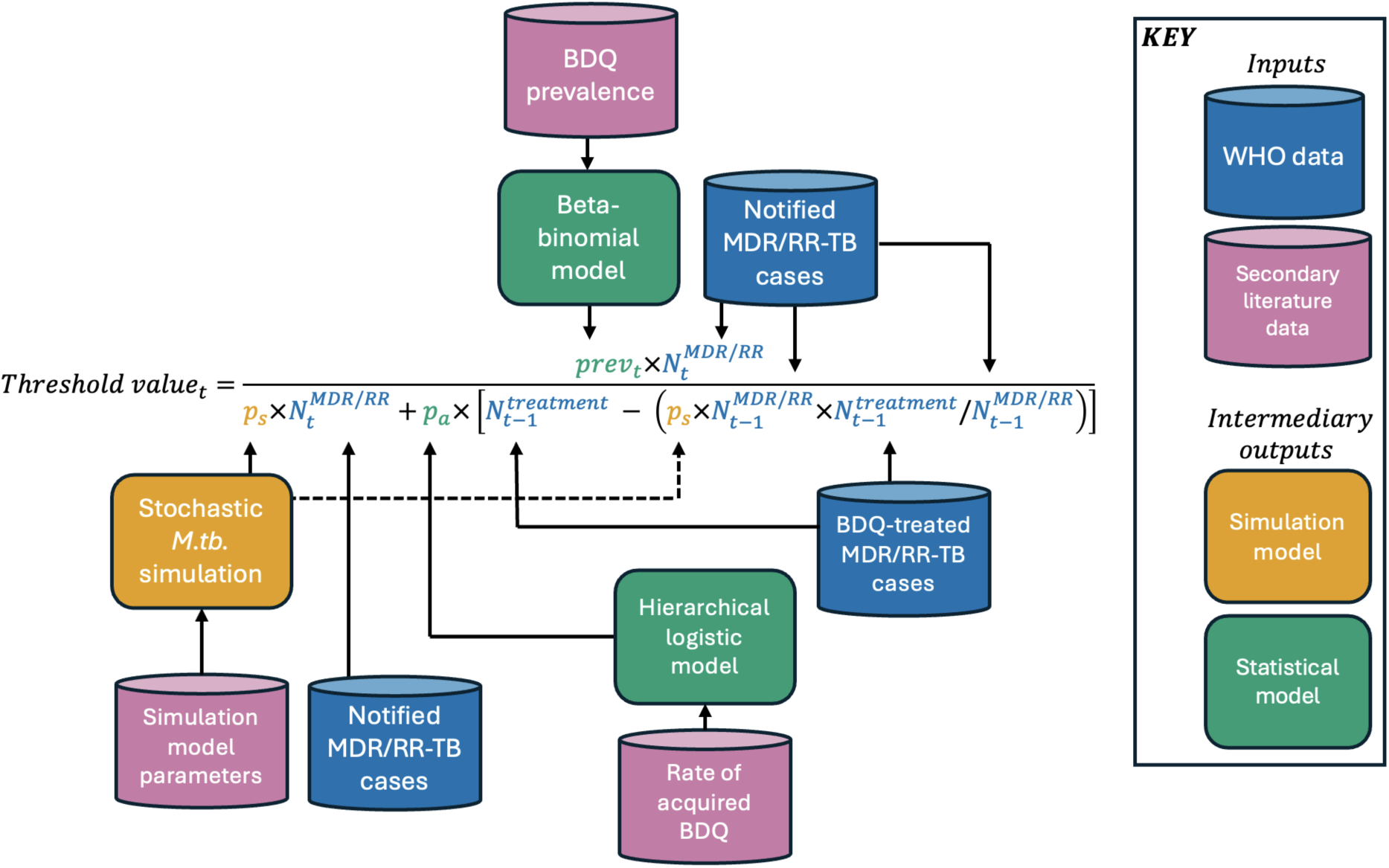
Transmission threshold model with data sources. WHO data (blue) are inputted directly into the model. BDQR prevalence and the probability of acquired resistance are drawn from posterior distributions of a beta-binomial model and a hierarchical logistic model (green) fitted to data from the literature (pink). The probability of spontaneous resistance is estimated from a simulation model (yellow) parameterised by data from the literature (pink). Dashed arrow for legibility purpose.

As we sought to estimate the relative contribution of transmission, we omitted directly quantifying 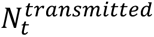 in (eq. 1). Instead, we devised the following transmission threshold value—the ratio of observed to expected—to answer when spontaneous and acquired resistance account (or, fail to account) for all observed resistance:

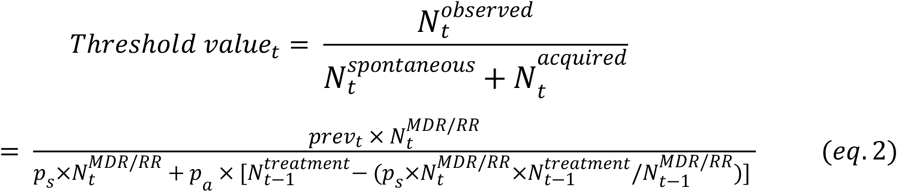

### Scenario analysis

To establish model behavior and determine the relationship between BDQ use (acquisition) and transmission, we conducted scenario analyses across a broad range of plausible parameter values, reflective of expanding BDQ roll-out and increasing BDQR burden. We varied the percentage of MDR/RR-TB patients treated with BDQ in the previous year between 50%-90% and the percentage of BDQR among MDR/RR-TB between 1%-5%.

### Country-level estimates

For each country-year, the threshold model was run 10,000 times, drawing samples from the random variables with associated uncertainty (*p*_*s*_, *p*_*a*_, and *prev*). The median, 2.5%, and 97.5% percentiles were reported. A *Threshold value*_*t*_ > 1 implies that considering spontaneous emergence and acquired resistance alone insufficiently account for observed levels of BDQR among MDR/RR-TB cases, and hence, that transmission may be occurring. *Threshold value*_*t*_ = 1 implies that the observed BDQR is fully accounted for by spontaneous and acquired resistance mechanisms—expected in settings where BDQ is still being rolled out. Finally, *Threshold value*_*t*_ < 1 implies that observed BDQR levels are below what would be expected from spontaneous and acquired resistance alone.

### Sensitivity analyses of country-level estimates

To explore the robustness of our results to underlying assumptions, we conducted the following one-way sensitivity analyses on the country-level estimates. First, we varied key random parameters (*prev*, *p*_*s*_, and *p*_*a*_) (Table 1) by a factor of two from their baseline values. For simplicity, we varied the probability of spontaneous resistance (*p*_*s*_) directly rather than the inputs to the simulation of this parameter—such as the limit of detection, critical bacteria population size, or per-division BDQR mutation rate. Second, we substituted WHO estimated MDR/RR-TB for notified MDR/RR-TB. Although the primary ambition was to assess whether evidence of transmission could be approached from readily observable data—which we expect to be more relevant for policymakers—we assessed the model’s sensitivity to potential underreporting that may be seen in notified cases. Moreover, we assessed the impact of potentially underreported treatment, by multiplying the number of MDR/RR-TB receiving BDQ by the ratio of estimated to notified MDR/RR-TB.

## Results

### Parameter estimates

We estimated the percentage of MDR/RR-TB cases spontaneously developing BDQR without BDQ treatment (*p*_*s*_) to be 0.073% (95% CrI: 0.062-0.085%). The mean probability of acquired BDQR (*p*_*a*_) was calculated as 3.6% (95% CrI: 2.8-4.4%). 18 studies were included through the systematic review, providing data on BDQR prevalence among MDR/RR-TB patients in 18 countries (Figure 2). BDQR prevalence among MDR/RR-TB patients ranged between 0-20% with study population sizes ranging from 1 to 2,308.

**Figure 2:**
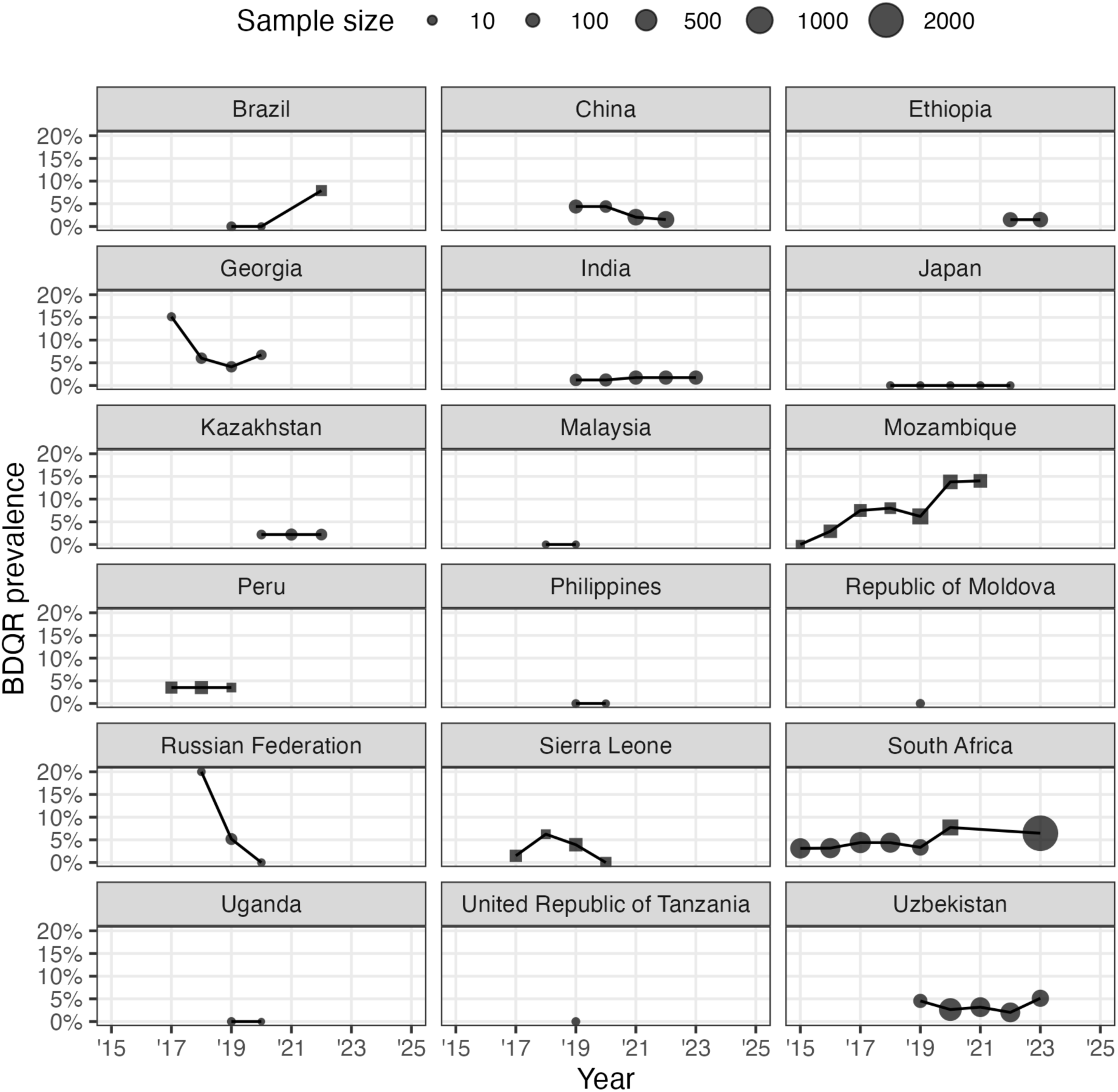
Annual national estimates of BDQR prevalence among MDR/RR-TB patients for 18 countries identified through systematic review. For each country (panel), the lines connect estimates from multiple years and studies, with the size of points indicating the sample size underlying the estimated BDQR prevalence. Squares are results from genotypic drug-susceptibility testing. Circles are results from phenotypic drug-susceptibility testing.

### Scenario analysis

The relative contribution of transmission to BDQR levels among MDR/RR-TB depends on both BDQR prevalence among MDR/RR-TB and the percentage of the MDR/RR-TB treated with BDQ (Figure 3). For a given year, the model suggest that transmission is likely occurring if BDQR prevalence among MDR/RR-TB exceeds 1.9% given that ≤50% of MDR/RR-TB patients are treated with BDQ in the preceding year (Figure 3). At low treatment rates (x-axis, Figure 3), and therefore few acquired cases, modest BDQR prevalence levels (y-axis) are sufficient to suggest transmission among MDR/RR-TB patients. As treatment rates increase, however, a proportional increase in BDQR prevalence is required to keep the threshold above 1 (dashed line, Figure 3).

**Figure 3:**
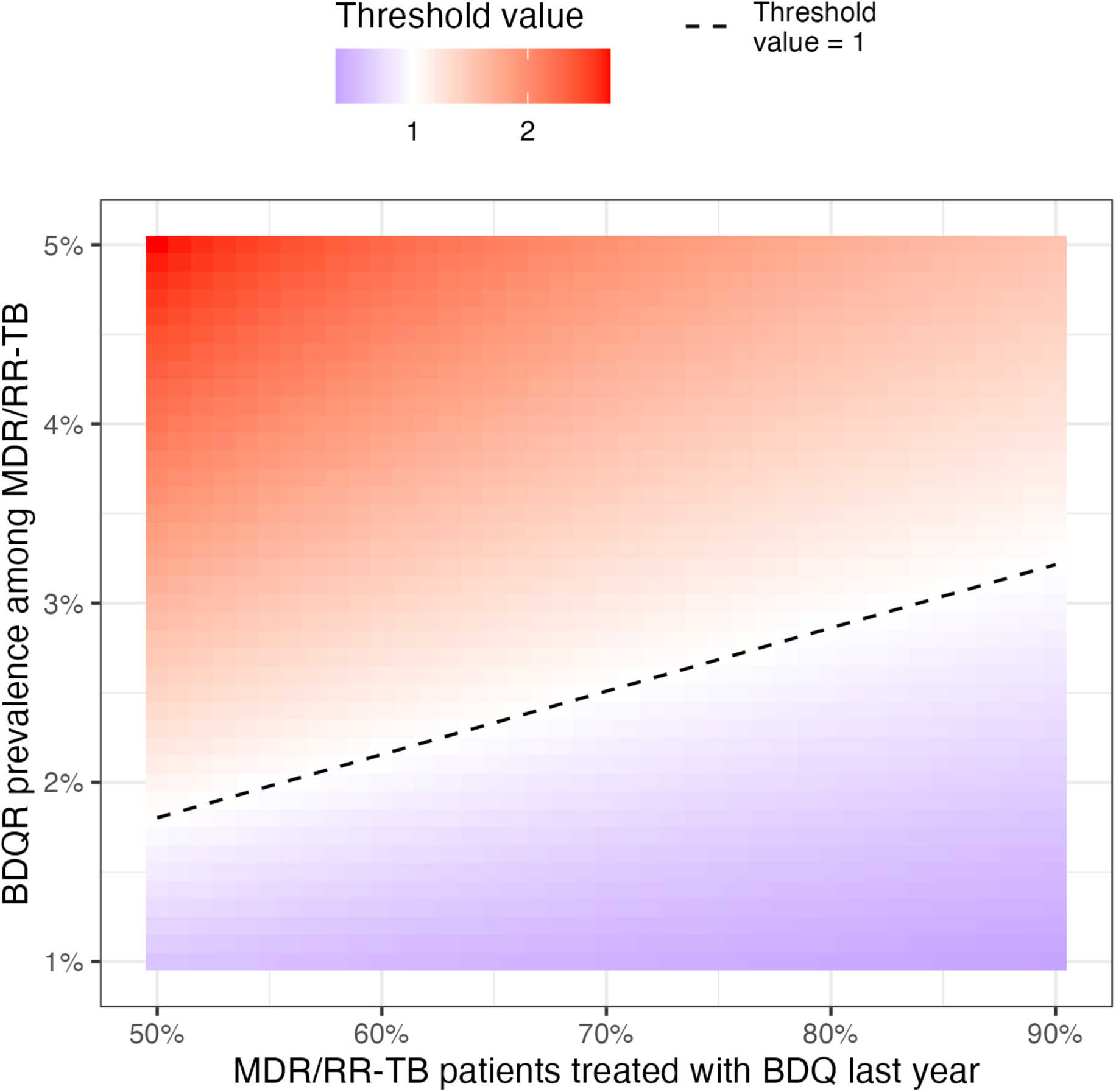
Transmission threshold depends on both the percentage of MDR/RR-TB patients treated with BDQ last year (x-axis) and BDQR prevalence among MDR/RR-TB patients (y-axis). Transmission must be invoked (red colours) when BDQR prevalence is high and treatment rates are low. The colour shows the estimated threshold value using baseline parameters (Table 1) and the dashed line the switchover point to requiring transmission to explain BDQR levels (threshold > 1).

### Country-level transmission thresholds

The model highlights likely variation in transmission dynamics across the 18 included countries that had BDQR prevalence data (Figure 4). Four countries (Brazil, China, Mozambique, and South Africa) had sufficient data to show periods of relatively stable transmission threshold values and associated 95% credible intervals >1 from 2020 onwards, indicating that BR-MDR/RR-TB burden exceeded the levels expected from spontaneous and acquired cases alone, and hence, suggesting possible BR-MDR/RR-TB transmission (Figure 4).

**Figure 4:**
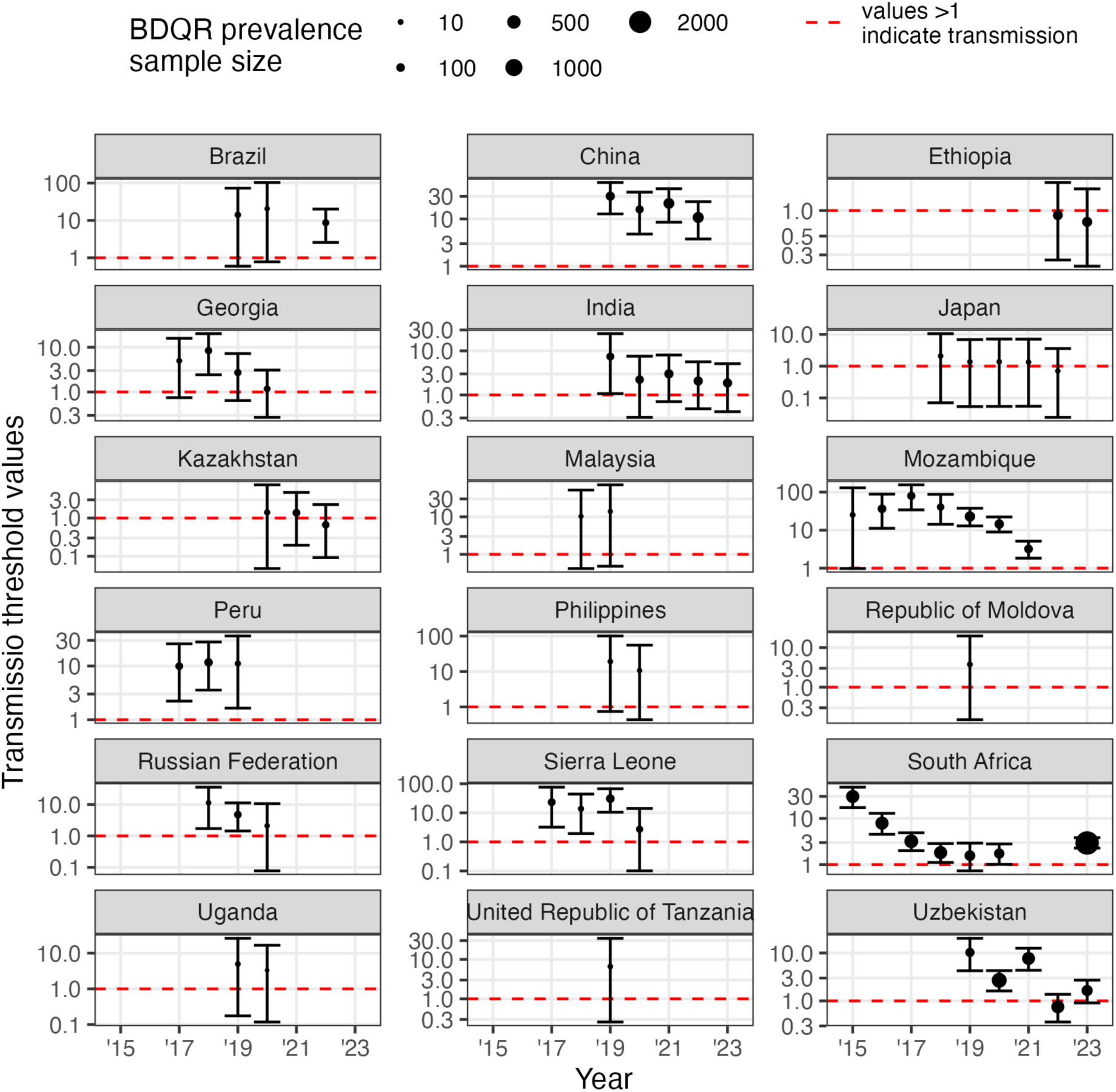
Threshold of transmission values by country and year. Points are median estimates over 10,000 simulations. Error bars are 95% credible intervals. Horizontal red dashed line = 1; if the threshold value is above 1, transmission is predicted to be contributing to the BDQR prevalence. Point sizes are proportional to sample size used to calculate BDQR prevalence. Note, y-axes differ between subplots and are shown on a logarithmic scale.

The relationship between treatment and BDQR prevalence seen in the scenario analysis (Figure 3), is reflected in the country level analysis (Figure 4). Although there is substantial heterogeneity and uncertainty, periods of declining transmission threshold values across most countries are apparent. In three countries (India, South Africa, and Mozambique), this downward trajectory occurred despite an increase in the BDQR prevalence among MDR/RR-TB over time. This is driven by proportionally faster increasing BDQ treatment rates over time, increasing the expected number of acquired resistant cases in the model’s denominator (corresponding to a horizontal shift in Figure 3). In India and Mozambique, although the relative transmission intensity decreased over time (lower threshold values), the absolute BR-MDR/RR-TB increased: from 1,186 to 1,600 between 2019-2023 in India and from 59 to 197 between 2015-2021 in Mozambique. In South Africa, the absolute burden of BR-MDR/RR-TB decreased over time from 680 to 570 between 2015-2023, driven in part by a more than 50% decrease in annual MDR/RR-TB incidence over the same period.

Three countries (Ethiopia, Japan, Kazakhstan) had transmission threshold values mostly near 1, indicating that observed BDQR prevalence is accounted for by the expected contribution of acquired and spontaneous resistance—as expected in countries where BDQ is still being rolled out and substantial transmission is not occurring.

### Sensitivity analysis

Results were generally robust to substantial variation in model parameters. Varying the probability of spontaneous resistance (*p*_*s*_) changed the qualitative interpretation in tree country-years towards transmission when halved and in one country-years away from transmission when doubled, based on the lower bound of 95% credible intervals being above or below 1— underscoring the small contribution of this mechanism to national BDQR burden (Supplement S6).

Varying the probability of acquired resistance (*p*_*a*_) and BDQR prevalence (*prev*)— corresponding to a horizontal and vertical shift in Figure 3 respectively—had a comparatively larger impact. However, qualitative interpretations based on the lower bound of 95% credible intervals only changed in country-years that were “edge cases”. Doubling *prev* shifted qualitative interpretations towards transmission in ten country-years—the same impact was observed in five of those ten country-years when halving *p*_*a*_. Halving *prev* shifted qualitative interpretations away from transmission in eight country-years—the same impact was observed in those same eight country-years when doubling *p*_*a*_ (Supplement S6).

For all countries and years except South Africa 2016, WHO estimated MDR/RR-TB was larger than notified. Substituting estimated for notified MDR/RR-TB shifted the qualitative interpretation towards transmission in three country-years. Finally, scaling BDQ treatment of MDR/RR-TB patients by the same ratio of estimated to notified MDR/RR-TB—generally corresponding to a positive horizontal shift in Figure3—shifted the qualitative interpretation away from transmission in six country-years.

## Discussion

In this study, we developed a transmission threshold model to give annual and country-specific quantitative evidence on the occurrence of BR-MDR/RR-TB transmission, by estimating where spontaneous and acquired resistance fail to account for observed BDQR prevalence among MDR/RR-TB populations. To parameterise the model, we calculated updated estimates of spontaneous and acquired BDQ resistance probabilities, and conducted a systematic review of BDQR prevalence studies, identifying 18 studies with data on 18 countries. Noticeably, our updated probability of acquired BDQR while on treatment of 3.6%—which is based on the most recently available data—is substantially higher than previous estimates of 2.1%-2.2%, suggesting acquired BDQR may be an even bigger concern than previously assumed [12,13]. This estimate may be particularly useful as a parameter for future modelling, as well as an expectation of outcomes for future treatment cohorts.

The primary finding from our modelling suggested that transmission may be required as an explanatory mechanism when BDQR prevalence among MDR/RR-TB cases exceeds 1.9% in settings where ≤ 50% of MDR/RR-TB patients were treated with BDQ in the preceding year. As BDQ use increases, for example to cover 90% of MDR/RR-TB patients, the prevalence of BDQR only has to increase to 3.3% to still suggest transmission (Figure 3). These threshold values provide important insights to the drivers of BR-MDR/RR-TB epidemiology, and indications for policymakers to shift intervention focus from adherence support to transmission interruption.

Some countries appeared to exhibit downward trends in threshold values, suggesting that later years were associated with relatively less transmission, despite BDQR prevalence being stable or even increasing. This paradoxical trend is driven by BDQ use expanding faster than BDQR prevalence, reducing the relative contribution of transmission. Rather than implying a true decline in transmission, this pattern may reflect countries’ transition through stages likely of any new antibiotic: early sporadic hotspot transmission, which may be oversurveilled, subsequent accumulation of resistance through expanding antibiotic use, and eventual endemic national transmission.

For example, in South Africa, Mozambique, and India, early observations suggest intense transmission, however, this pertains to few absolute BDQR cases, which are unexplainable by low BDQ treatment rates. These observations may be consistent with isolated outbreaks such as nosocomial transmission, or unreliable early surveillance, when BDQR testing standards and coverage were inconsistent and BDQ was prescribed to patients with high pre-existing resistance. While BDQR prevalence increases, BDQ use grows more rapidly, lowering our calculated threshold values—potentially because BDQ use may extend to populations with less pre-existing resistance, hence, overestimating the contribution of this mechanism, or because effective BDQ use decreases the pool of individuals transmitting BR-MDR/RR-TB over time. In South Africa, an early and extensive adopter of BDQ, the BDQ treatment rate has practically been saturated post 2020, meaning continuing increases in BDQR prevalence leads to increasing relative transmission in the model. This may indicate endemic transmission, as although threshold values are lower in 2023 than 2015, this transmission pertains to a much larger proportion of all MDR/RR-TB being BDQR (6.5% vs 3.2%).

In several country-years (Ethiopia (2022-2023), Japan (2022), Kazakhstan (2022), and Uzbekistan (2022)), point threshold values were below one, suggesting fewer BR-MDR/RR-TB cases than expected from spontaneous and acquired resistance. While this may be evidence against transmission occurring, it may also indicate data limitations. For example, underreporting of MDR/RR-TB would negatively bias threshold values: our sensitivity analysis showed that for these three countries, substituting notified for reported MDR/RR-TB, shifted the threshold point values above one for Ethiopia, Japan, and Uzbekistan (Supplement S6). Crucially, this highlights that modelled results may underestimate transmission in cases where MDR/RR-TB is substantially underdetected and/or underreported.

The model was robust to large variation in the probability of spontaneous mutation, underscoring the small impact of this mechanism. Variation in the probability of acquired resistance impacted qualitative interpretations for country-level on several occasions, underscoring the potential epidemiological benefit of managing acquired resistance during expanding access to new antibiotics.

Previous modelling by Kendall et al. showed that for established anti-TB drugs, namely rifampicin and isoniazid, the overwhelming majority of resistance (up to >99% in some contexts) is transmitted [7]. Our work complements their approach by focusing on the early stage of antibiotic introduction and the transition point from acquisition to transmission predominance. While their analysis did not consider spontaneous resistance, we show that the impact of this mechanism is likely miniscule compared to acquired resistance.

Our study is not without limitations. First, BDQR data is limited and to our knowledge no nationally representative BDQR prevalence survey has been conducted. To fill this gap, we conducted a systematic review and assumed extrapolation from identified studies to the national level was possible. There were substantial differences in sample sizes, study designs, and assumptions about the representativeness of BDQR prevalence data may be particularly weak in historical (pre-2020) data. We address this uncertainty by including it in our credible intervals and by varying BDQR prevalence in the sensitivity analysis. Second, the model assumes a closed population. In some settings, BR-MDR/RR-TB may in part be driven by population movements: for example, migration between Mozambique and South Africa may impact BDQR burdens in both countries [14]. Finally, we did not model cross-resistance (to clofazimine) explicitly because of limited data. This mechanism is partly captured within our spontaneous resistance parameter—which we inflate during sensitivity analysis. Acquired and spontaneous resistance may be underestimated in early timepoints as BDQ was initially used mainly in hard-to-treat, potentially more BDQR-prone, patients and background clofazimine use would be more important for BDQR cross-resistance before BDQ use became widespread. We addressed this in the sensitivity analysis, which did not impact qualitative country-level interpretations in early timepoints.

This study has several important strengths. We combine multiple data sources, including our systematic review of BDQR prevalence among MDR/RR-TB, and provide renewed estimates of acquired and spontaneous resistance. Our parsimonious model also allowed for better understanding of the impact of constituent model components, as well as retaining the uncertainty in each model component in the ultimate output.

Our study has several implications for future research. First, the model’s predominant reliance on readily available notification data makes it possible to update estimates as new data becomes available—including in countries not analysed in this study—and could serve as a first modelled indication of BDQR transmission or transmission of resistance to other new TB antibiotics. Future research generating representative longitudinal data on BDQR prevalence would allow further explorations of temporal dynamics and confidence in the timings of transition towards transmission dominance. Second, our model is generalisable to other TB drugs and bacterial infection—although, the *M.tb.* simulation is specifically made for a strictly clonal pathogen, and hence, may not be well suited for pathogens with significant horizontal gene transfer.

In conclusion, we present a novel tool for assessing mechanisms for resistance in the early stages of new drug usage for TB, providing new key parameter estimates and evidence of BDQR transmission. Understanding the relative contribution of spontaneous resistance, acquisition, and transmission at a given timepoint may help guide whether programmatic focus should be on reducing acquisition or transmission interruption.

## Data Availability

All data and code supporting the manuscript have shared publicly to a GitHub repository.

https://doi.org/10.5281/zenodo.18210207

## Acknowledgement

The authors would like to thank Theolis Barbosa, Popy Yuniar, Nugroho Soeharno, and Sarika Mehra for their valuable feedback on this work.

## References

1. World Health Organization. WHO consolidated guidelines on tuberculosis Module 4: Treatment Drug-resistant tuberculosis treatment. World Health Organization; 2022. Available: https://iris.who.int/server/api/core/bitstreams/b4112461-7e9c-403c-808f-055f1dc3a54b/content

2. World Health Organization. Global tuberculosis report 2024. World Health Organization; 2024. Available: https://www.who.int/teams/global-programme-on-tuberculosis-and-lung-health/tb-reports/global-tuberculosis-report-2024

3. Weiner B, Gomez J, Victor TC, Warren RM, Sloutsky A, Plikaytis BB, et al. Independent Large Scale Duplications in Multiple M. tuberculosis Lineages Overlapping the Same Genomic Region. Tailleux L, editor. PLoS ONE. 2012;7: e26038. doi:10.1371/journal.pone.0026038

4. Hirsh AE, Tsolaki AG, DeRiemer K, Feldman MW, Small PM. Stable association between strains of *Mycobacterium tuberculosis* and their human host populations. Proc Natl Acad Sci. 2004;101: 4871–4876. doi:10.1073/pnas.0305627101

5. Mallick JS, Nair P, Abbew ET, Van Deun A, Decroo T. Acquired bedaquiline resistance during the treatment of drug-resistant tuberculosis: a systematic review. JAC-Antimicrob Resist. 2022;4: dlac029. doi:10.1093/jacamr/dlac029

6. Hu X, Wu Z, Lei J, Zhu Y, Gao J. Prevalence of bedaquiline resistance in patients with drug-resistant tuberculosis: a systematic review and meta-analysis. BMC Infect Dis. 2025;25: 689. doi:10.1186/s12879-025-11067-2

7. Kendall EA, Fofana MO, Dowdy DW. Burden of transmitted multidrug resistance in epidemics of tuberculosis: a transmission modelling analysis. Lancet Respir Med. 2015;3: 963–972. doi:10.1016/S2213-2600(15)00458-0

8. Nguyen TVA, Anthony RM, Bañuls A-L, Nguyen TVA, Vu DH, Alffenaar J-WC. Bedaquiline Resistance: Its Emergence, Mechanism, and Prevention. Clin Infect Dis. 2018;66: 1625–1630. doi:10.1093/cid/cix992

9. Ismail N, Sirgel F, Omar SV, Omar S, De Kock M, Spies C, et al. Unpacking bedaquiline heteroresistance: the importance of intermediate profiles for phenotypic drug susceptibility testing. Shields RK, editor. Antimicrob Agents Chemother. 2025; e00356–25. doi:10.1128/aac.00356-25

10. Degiacomi G, Sammartino JC, Sinigiani V, Marra P, Urbani A, Pasca MR. In vitro Study of Bedaquiline Resistance in Mycobacterium tuberculosis Multi-Drug Resistant Clinical Isolates. Front Microbiol. 2020;11: 559469. doi:10.3389/fmicb.2020.559469

11. Andries K, Villellas C, Coeck N, Thys K, Gevers T, Vranckx L, et al. Acquired Resistance of Mycobacterium tuberculosis to Bedaquiline. Van Veen HW, editor. PLoS ONE. 2014;9: e102135. doi:10.1371/journal.pone.0102135

12. Mallick JS, Nair P, Abbew ET, Van Deun A, Decroo T. Acquired bedaquiline resistance during the treatment of drug-resistant tuberculosis: a systematic review. JAC-Antimicrob Resist. 2022;4: dlac029. doi:10.1093/jacamr/dlac029

13. Perumal R, Bionghi N, Nimmo C, Letsoalo M, Cummings MJ, Hopson M, et al. Baseline and treatment-emergent bedaquiline resistance in drug-resistant tuberculosis: a systematic review and meta-analysis. Eur Respir J. 2023;62: 2300639. doi:10.1183/13993003.00639-2023

14. Barwise K, Lind A, Bennett R, Martins E. Intensifying Action to Address HIV and Tuberculosis in Mozambique’s Cross-Border Mining Sector. Int J Health Serv. 2013;43: 699–719. doi:10.2190/HS.43.4.g

