## Supplement for "Estimating the relative contribution of transmission to bedaquiline resistance burden in tuberculosis. A transmission threshold model"

#### S1 Calculating probability of spontaneous resistance

We estimated $p_s$, the probability of spontaneous resistance, using a within-host *M.tb.* simulation, adapted from Colijn et al. [1]. We simulated the stochastic growth and emergence of BDQ resistance through *de-novo* mutations in an initially fully BDQ susceptible *M.tb.* population.

In the model, we assumed an initial BDQ-susceptible *M.tb.* population of 10, $N(0) = 10$, and BDQ-resistant population of 0, $M(0) = 0$. While the bacteria population is relatively small, $N < 10{,}000$, a monte carlo simulation was used to model $N(t+1)$: at every timestep, $t$, for bacteria $i = 1,2,\ldots,N(t)$, the $ith$ BDQ-susceptible *M.tb.* bacteria divides with probability $\lambda$. If the bacteria divides, the offspring is a BDQ-resistant mutant with probability $\beta$. At the same timestep, parent bacteria dies with probability $\mu$. The BDQ-resistant population was modelled similarly with division probability $\lambda_1$ and death probability $\mu_1$.

Due to the computational demand of the monte carlo approach, when the bacteria population reached a large size, $N = 10{,}000$, we approximated $N(t+1) = N(t) + divisions_N - mutations - deaths_N$, where $divisions_N$, $deaths_N$, and $mutations$ were binomially distributed with parameters $Binomial(N(t), \lambda)$, $Binomial(N(t), \mu)$, $Binomial(divisions, \beta)$ respectively. Similarly, $M(t+1) = M(t) + divisions_M - deaths_M + mutations$, where $divisions_M$ and $deaths_M$ were binomially distributed with parameters $Binomial(M(t), \lambda_1)$ and $Binomial(M(t), \mu_1)$ respectively.

We assumed the per-division probability of a BDQR-conferring mutation was $5 \times 10^{-7}$—the upper bound of estimates from *in vitro* studies [2]—and no fitness cost for the BDQR strain [3,4]. We assumed that detection and sputum collection would occur when the bacteria population reached $10^{10}$. Due to the simulation being implemented in discrete time, we simulated each bacteria population until $N(t) \geq 1.1 \times 10^{10}$ and linearly interpolate the time ($\tau$) at which $N(t = \tau) = 10^{10}$. We then approximated the resistant bacteria population at this time by using the same interpolation $M(t = \tau)$. We independently simulated 200,000 populations and collected the proportion of simulations in which $M(t = \tau)/N(t = \tau) \geq 1\%$—we assumed BDQR was detectable if $\geq 1\%$ of the bacteria population was resistant, reflecting the limit of detection for phenotypic DST in heteroresistant bacterial populations [5]. We then fitted a Bayesian beta-binomial model with a flat prior to the proportion of simulated bacteria populations where resistance was detectable. $p_s$ was sampled from the resultant posterior distribution.

The model was implemented with a daily timestep and assumed a daily cell-cycle ($\lambda = 1$). We calculated the death probability ($\mu$) from the exponential approximation:

$$N_\tau = N_0 \times exp[(\lambda - \mu) \times \tau)] \qquad (eq.S1)$$

, where $N_\tau = 10^{10}$, $N_0 = 10$, $\lambda = 1$, and $\tau = 1$ year. The full parameter set used to estimate $p_s$ are presented in Table S1.

| Symbol | Description | Value | Source |
|---|---|---|---|
| $\lambda$ | Probability of bacteria division. | 1 | assumed |
| $\mu$ | Probability of bacteria death. | 0.94 | Calculated |
| $\lambda_1$ | Probability of bacteria division, BDQ-resistant strain. | 1 | Andries et al. [4] |
| $\mu_1$ | Probability of bacteria death, BDQ-resistant strain. | $\mu$ | Andries et al. [4] |
| $\beta$ | Probability of spontaneous BDQ-resistance mutation. | $5 \times 10^{-7}$ | Nguyen et al. [2] |
| $N_\tau$ | Size of bacteria population when host seeks treatment and BDQ-resistance is evaluated | $10^{10}$ | Colijn et al. [6] |
| | Phenotypic drug susceptibility limit of detection | 1% | Ismail et al. [5] |

**Table S1:** simulation parameters for estimation of probability of spontaneous resistance.

### S2 Calculating probability of acquired resistance

We estimated $p_a$, the probability of acquiring resistance to BDQ during treatment, using pooled data from two systematic reviews. We combine data from 11 studies from a 2022 review by Mallick et al., comprising 1,087 individuals, and a further 5 studies from a 2025 review by Hu et al., comprising 1,056 individuals [7,8]. The final sample contained 2,143 individuals from 16 studies. We extracted the number of acquired BDQR cases ($x_i$) and the total number of patients ($n_i$) from each study ($i \in 1,2,\ldots,N$). Then, we fitted a Bayesian hierarchical logistic model to the data to account for between-study heterogeneity. We assumed that the log-odds of acquired bedaquiline resistance varied between studies with a normally distributed shared mean. Hence we have:

$$x_i \sim Binomial(n_i, p_i) \qquad (eq.S2)$$

With $p_i = logit^{-1}\theta_i$. $\theta_i$ is the log-odds of acquired bedaquiline resistance in study $i$ , which we assume is normally distributed with a shared population mean $\mu$ and standard deviation $\tau$, hence:

$$\theta_i \sim Normal(\mu, \tau^2) \qquad (eq.S3)$$

With weakly informative priors (note that $\mu$ is on the log-odds scale): $\mu \sim Normal(-2.5, 1)$ and $\tau \sim Cauchy(0, 1)$

We then collect $p_a$ as the weighted mean posterior distribution of $p_i$ across studies:

$$p_a = \frac{1}{\sum_i^N \quad n_i} \times \sum_i^N \quad (n_i \times logit^{-1}\theta_i) \qquad (eq.S4)$$

### S3 Calculating prevalence of BDQR among MDR/RR-TB

In order to identify studies reporting the prevalence of BDQR among MDR/RR-TB cases, we searched PubMed on 26 June 2025 using the following string:

"(bedaquiline[Title/Abstract] **OR** TMC207[Title/Abstract]) **AND**
(resistance[Title/Abstract] **OR** resistant[Title/Abstract]) **AND**
(prevalence[Title/Abstract] **OR** baseline[Title/Abstract]) **AND**
(tuberculosis[MeSH Terms] **OR** tuberculosis[Title/Abstract] **OR** TB[Title/Abstract])"

The inclusion and exclusion discussed in the main text. Study summaries are presented below (Table S2).

For each country year, where prevalence data was identified, we fitted a Bayesian beta-binomial model to the calculated prevalence and sample size. We then draw posterior samples from the fitted model as $prev$. To stabilise inference in country-years with small sample sizes, we used a weakly informative Beta prior centered on low resistance prevalence, reflecting epidemiological plausibility.

| Reference | ISO | Study overview | Data cleaning / year imputation | Considerations |
|---|---|---|---|---|
| Barilar (2024) | MOZ | Source: National TB Reference lab<br>Test: genotypic<br>Location: Mozambique<br>When: 2015-2021<br>Population: RR-TB (WGS) | Aggregate: 61/704 (8.7% ) were BDQR<br>Cleaning: individual level data reported. | XDR is defined as pre-XDR + resistance to combined fluoroquinolone and bedaquiline resistance (in abstract); as pre-XDR + resistance to at least one of additional Group A drugs bedaquiline or linezolid (main text). Assuming all XDR were BDQR matched prevalences reported by the authors in the main manuscript. |
| Blankson (2024) | SLE | Source: National TB reference lab<br>Test: genotypic<br>Location: Freetown, Sierra Leone<br>When: Jan 1, 2015 - July 31, 2021<br>Population: RR-TB (Xpert) or suspected RR-TB + failed first-line treatment | Aggregate: 6/235 (2.6%) were BDQR<br>Cleaning: individual level data reported. | |
| Diriba (2025) | ETH | Source: National TB reference lab.<br>Test: phenotypic<br>Location: Ethiopia<br>When: February 2022 - July 2024<br>Population: MDR/RR-TB | Aggregate: 7/468 (1.5%) were BDQR<br>Cleaning: BDQR prevalence was disaggregated using imputation* | |
| Hu (2023) | CHN | Source: Chongqing Tuberculosis Control Institute<br>Test: phenotypic<br>Location: Chongqing<br>When: March 2019 - June 2020<br>Population: MDR-TB with suggestive active TB. | Aggregate: 9/205 (4.4%) were BDQR.<br>Cleaning: BDQR prevalence was disaggregated using imputation* | |

| | | | | |
|---|---|---|---|---|
| Ismail (2022) | ZAF | Source: National reference lab (NICD)<br>Test: phenotypic<br>Location: South Africa<br>When: Jan 1, 2015 - July 31, 2019<br>Population: all TB patients starting BDQ treatment (at least RR) | Aggregate: 76/2023 (3.8%) were BDQR.<br>Cleaning: the authors reported samples per guideline period rather than year. We assumed uniform distribution of samples and BDQR risk throughout guideline periods and disaggregated using imputation *. | |
| Li (2024) | CHN | Source: Four TB specialized hospitals<br>Test: phenotypic<br>Location: four provinces in China<br>When: 2022<br>Population: MDR-TB | Aggregate: 4/263 (1.5%) were BDQR<br>Cleaning: all samples were from a single year | |
| Marcon (2025) | BRA | Source: National reference lab<br>Test: genotypic<br>Location: State of Pará<br>When: 2021-2022<br>Population: DR-TB | Aggregate: 3/38 were BDQR<br>Cleaning: BDQR prevalence was disaggregated using imputation* | Samples were collected between November 2021 - December 2022. Only two samples were from 2021 and were both BDQS. We excluded these and extracted data from 2022 only. |
| Mikiashvil (2024) | GEO | Source: Retrospective programmatic setting<br>Test: phenotypic<br>Location: Georgia<br>When: November 2017- December 2020.<br>Population: MDR/RR-TB treated with BDQ and delamanid. | Aggregate: 6/89 (6.7%) were BDQR<br>Cleaning: BDQR prevalence was disaggregated using imputation* | |
| Moe (2024) | UZB | Source: Retrospective lab<br>Test: phenotypic<br>Location: Karakalpakstan<br>When: 2019 - 2023<br>Population: TB patients who underwent pDST for SLDR (55% were MDR, 1.3% RR, 26 IR, 18% DS.) | Aggregate: 61/1813 (3.4%) were BDQR<br>Cleaning: annual prevalence was reported. | There were 429 and 1 patients with unknown BDQR status in 2019 and 2020 respectively, which was not clarified by the authors. These patients were assumed not BDQR, which produced prevalences similar to other years. |

| | | | | |
|---|---|---|---|---|
| Moultrie (2024) | ZAF | Source: National reference lab<br>Test: phenotypic<br>Location: Eastern Cape, Western Cape, KwaZulu Natal<br>When: 2023<br>Population: RR-TB | Aggregate: 149/2,308<br>Cleaning: all samples were from a single year | Data is unpublished, but available in the following presentation: https://knowledgehub.health.gov.za/system/files/2024-03/In-Session%20slides_BDQ%20resistance%20webinar.pdf |
| Padmapriyadarsini (2024) | IND | Source: Prospective clinical trial<br>Test: phenotypic<br>Location: 9 study sites in India<br>When: 2021 - 2023<br>Population: pre-XDR or treatment intolerant/nonresponsive MDR (BDQ naive) | Aggregate: 7/403 7 (1.7%) were BDQR<br>Cleaning: BDQR prevalence was disaggregated using imputation* | Note, 3 were excluded before treatment initiation. In total 7 had baseline resistance (DST results arriving after treatment initiation). We assume that the total number of baseline BDQR were 7. |
| Padmapriyadarsini (2022) | IND | Source: Prospective clinical trial<br>Test: phenotypic<br>Location: 5 sites in India<br>When: April 2019 - January 2021<br>Population: 165 MDR-TBFQ+ or/and MDR-TBSLI+ | Aggregate: 2/165 (1.2%) were BQDR<br>Cleaning: BDQR prevalence was disaggregated using imputation* | |
| Rashitov (2024) | KAZ | Source: prospective 5 public health facilities under operational research conditions<br>Test: phenotypic<br>Location: Kazakhstan<br>When: 22 Sep 2020 - 30 Sep 2022<br>Population: MDR/RR-TB starting BDQ treatment. | Aggregate: 3/137 (2.2%) were BDQR<br>Cleaning: BDQR prevalence was disaggregated using imputation* | |
| Roberts (2024) | ZAF | Source: National reference lab<br>Test: genotypic<br>Location: Gauteng and Western Cape<br>When: Jan 10, 2019 - July, 22, 2020<br>Population: MDR-TB | Aggregate: 32/371 were BDQR<br>Cleaning: individual level data reported | |
| Timm | BRA, | Source: multiple trials | Aggregate: 12/334 were BDQR | |

| | | | |
|---|---|---|---|
| (2023) | GEO,<br>MYS,<br>MDA,<br>PHL,<br>RUS,<br>ZAF,<br>TZA,<br>UGA | Test: phenotypic<br>Location: multiple locations<br>When: 2015 - 2020<br>Population: DR-TB | Cleaning: individual level data reported |
| Tong (2023) | CHN | Source: 19 municipal TB hospitals<br>Test: LJ<br>Location: Zhejiang province<br>When: 2021<br>Population: RR-TB | Aggregate: 5/245 (2.1%) were BDQR<br>Cleaning: all samples were from a single year |

**Table S2:** summaries of studies providing BDQR prevalence data used in transmission threshold model, identified through systematic review. * imputation: in some studies, only the aggregate number of BDQ were given. Hence, we assume that sampling and BDQR prevalence were uniformly distributed throughout the study period and assign samples and BDQR prevalence to each year weighted by the number of months sampling occurred in that year: n/(months of sampling in year × total months of sampling)

### S4 Input data and missing data imputation

The annual BDQR prevalence as well as the annual WHO notified MDR/RR-TB incidence are presented below (Table S3). The WHO-notified number of MDR/RR-TB cases treated with BDQ were missing for 12 years in seven countries. Details of missingness and imputation strategy is discussed in the main text. The imputed number of MDR/RR-TB cases treated with BDQR is presented below (Table S3, Figure S1).

| | | WHO notification data | | Data from systematic review | |
|---|---|---|---|---|---|
| **Country** | **Year** | **BDQ treated preceding year (imputed)** | **MDR/RR-TB incidence** | **BDQR (%)** | **BDQS (%)** |
| Brazil | 2019 | 8 | 1039 | 0 (0%) | 21 (100%) |
| | 2020 | 10 | 960 | 0 (0%) | 4 (100%) |
| | 2022 | 206 | 1239 | 3 (7.5%) | 37 (92.5%) |
| China | 2019 | 336 | 18246 | 5.6 (4.4%) | 122.5 (95.6%) |
| | 2020 | 866 | 21016 | 3.4 (4.4%) | 73.5 (95.6%) |
| | 2021 | 110 | 19860 | 5 (2%) | 240 (98%) |
| | 2022 | 326 | 16275 | 4 (1.5%) | 259 (98.5%) |
| Ethiopia | 2022 | 376 | 661 | 7 (1.5%) | 461 (98.5%) |
| | 2023 | (483.5) | 773 | 7 (1.5%) | 461 (98.5%) |
| Georgia | 2017 | (87.1) | 339 | 7 (7.5%) | 86 (92.5%) |
| | 2018 | 45 | 311 | 7 (6.4%) | 102 (93.6%) |
| | 2019 | 117 | 319 | 6 (5.6%) | 101 (94.4%) |
| | 2020 | 228 | 191 | 6 (6.7%) | 83 (93.3%) |
| India | 2019 | 2865 | 66255 | 2 (1.2%) | 163 (98.8%) |
| | 2020 | 5513 | 36395 | 2 (1.2%) | 163 (98.8%) |
| | 2021 | 10140 | 74426 | 7 (1.7%) | 396 (98.3%) |

| | | WHO notification data | | Data from systematic review | |
|---|---|---|---|---|---|
| Country | Year | BDQ treated preceding year (imputed) | MDR/RR-TB incidence | BDQR (%) | BDQS (%) |
| | 2022 | 16752 | 82267 | 7 (1.7%) | 396 (98.3%) |
| | 2023 | (18516.9) | 81052 | 7 (1.7%) | 396 (98.3%) |
| Japan | 2018 | (27.4) | 94 | 0 (0%) | 4.6 (100%) |
| | 2019 | 30 | 68 | 0 (0%) | 4.6 (100%) |
| | 2020 | (25.5) | 58 | 0 (0%) | 4.6 (100%) |
| | 2021 | 25 | 55 | 0 (0%) | 4.6 (100%) |
| | 2022 | 40 | 43 | 0 (0%) | 3.9 (100%) |
| Kazakhstan | 2020 | 1152 | 3696 | 0.4 (2.2%) | 16.1 (97.8%) |
| | 2021 | 1325 | 3776 | 1.4 (2.2%) | 64.3 (97.8%) |
| | 2022 | 4123 | 4767 | 1.2 (2.2%) | 53.6 (97.8%) |
| Malaysia | 2018 | (9.2) | 192 | 0 (0%) | 2 (100%) |
| | 2019 | 5 | 169 | 0 (0%) | 2 (100%) |
| Mozambique | 2015 | (0) | 646 | 0 (0%) | 9 (100%) |
| | 2016 | (0) | 911 | 3 (2.9%) | 100 (97.1%) |
| | 2017 | (0) | 861 | 6 (7.5%) | 74 (92.5%) |
| | 2018 | 24 | 1158 | 4 (8%) | 46 (92%) |
| | 2019 | 68 | 1388 | 13 (6.2%) | 197 (93.8%) |
| | 2020 | 241 | 1130 | 20 (13.8%) | 125 (86.2%) |
| | 2021 | 1374 | 1346 | 15 (14%) | 92 (86%) |
| Peru | 2017 | (98.6) | 1508 | 2.2 (3.5%) | 60 (96.5%) |
| | 2018 | 102 | 1942 | 3.3 (3.5%) | 90 (96.5%) |
| | 2019 | 150 | 1894 | 0.5 (3.5%) | 15 (96.5%) |
| Philippines | 2019 | 72 | 7492 | 0 (0%) | 6 (100%) |

| Country | Year | WHO notification data | | Data from systematic review | |
|---|---|---|---|---|---|
| | | **BDQ treated preceding year (imputed)** | **MDR/RR-TB incidence** | **BDQR (%)** | **BDQS (%)** |
| | 2020 | 347 | 8401 | 0 (0%) | 3 (100%) |
| Moldova | 2019 | 83 | 655 | 0 (0%) | 9 (100%) |
| Russia | 2018 | 3167 | 27438 | 1 (20%) | 4 (80%) |
| | 2019 | 6436 | 27207 | 3 (5.2%) | 55 (94.8%) |

**Table S3:** input data for transmission threshold value estimation.

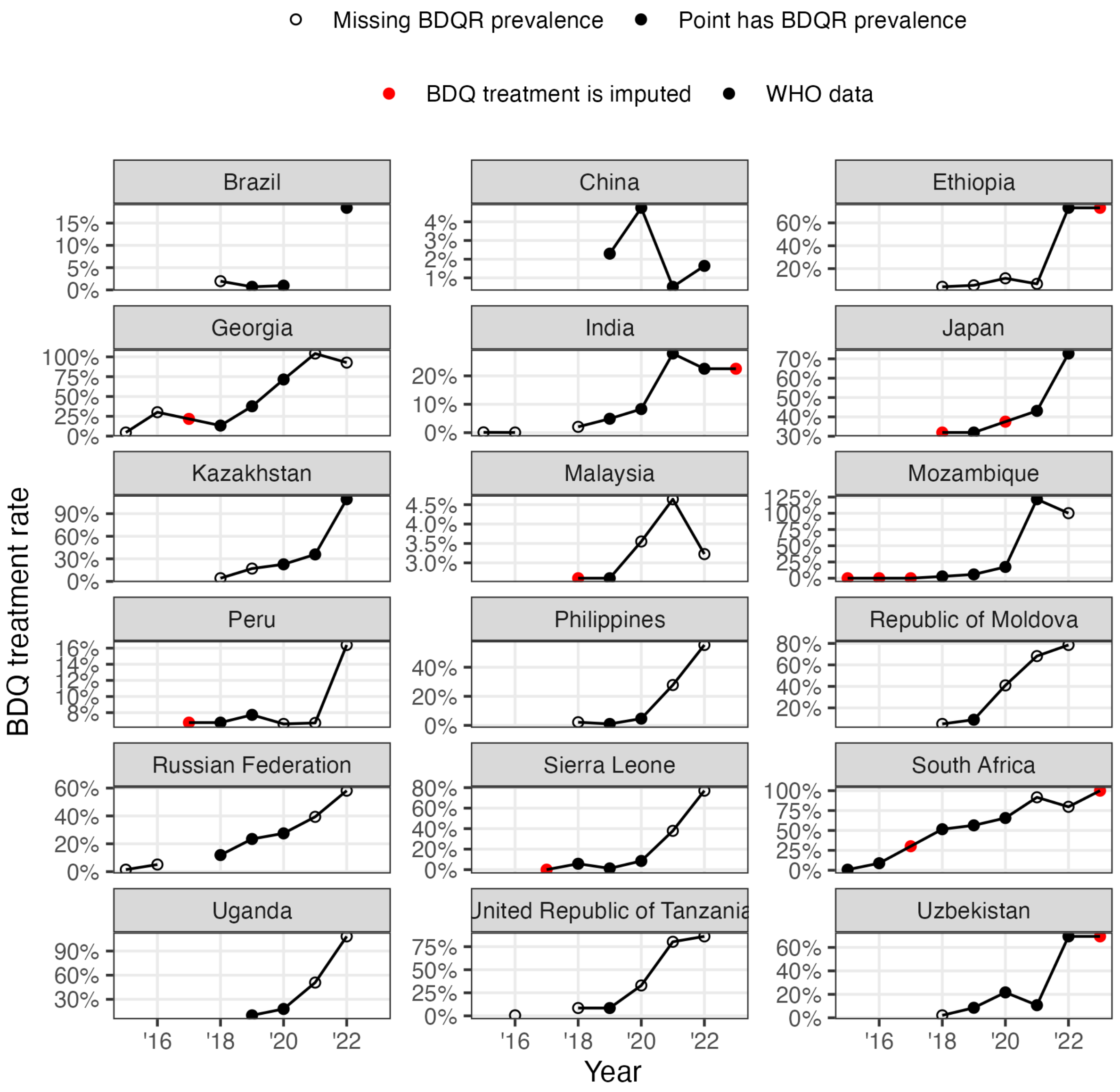


**Figure S1:** Annual WHO-notified BDQ treatment rate (percentage of incident MDR/RR-TB treated with BDQ). Highlighted in red are imputed treatment rates. Solid points are where BDQR prevalence was identified through systematic review (see main text) and threshold transmission value estimated. Hollow points are where BDQR prevalence was not identified and included only for visual context. Note, y-axes differ.

### S5 Treatment and BDQR prevalence impact on threshold estimates

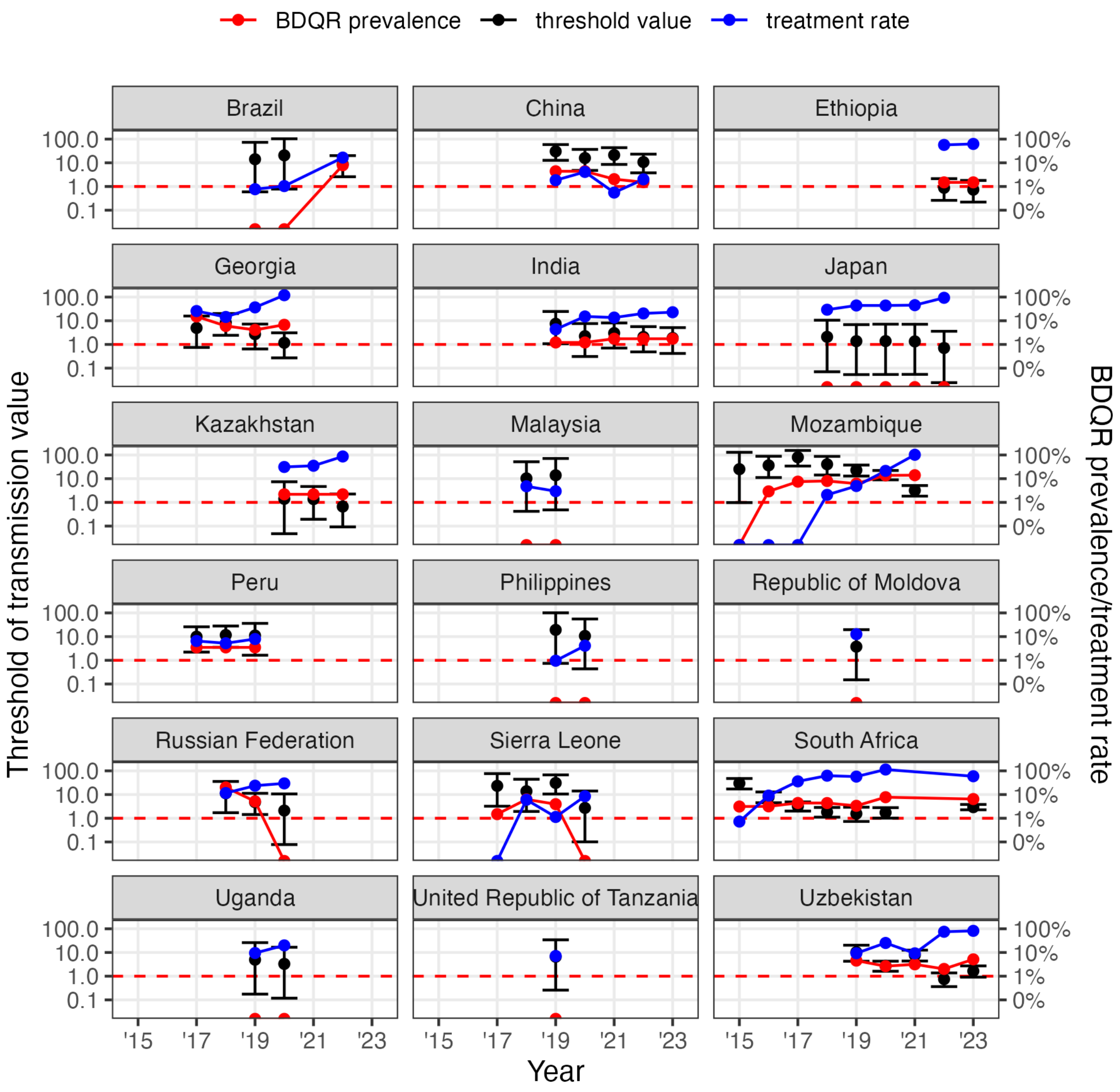


**Figure S2:** As BDQ use for MDR/RR-TB treatment increases, more acquisition of BDQR occurs, leading to a decrease in the transmission threshold values. Median threshold value (left y-axis). BDQR prevalence (red, %) and percentage of MDR/RR-TB cases treated with BDQ last year (blue, %) (right y-axis). Note, y-axes differ between subplots and shown on a logarithmic scale.

### S6 Sensitivity analysis

To explore the robustness of our results to underlying assumptions, we conducted the following one-way sensitivity analyses on the country-level estimates. First, we varied key random parameters ($prev$, $p_s$, and $p_a$) by a factor of two from their baseline values. For simplicity, we varied the probability of spontaneous resistance ($p_s$) directly rather than the inputs to the simulation of this parameter—such as the limit of detection, critical bacteria population size, or per-division BDQR mutation rate. Second, we substituted WHO estimated MDR/RR-TB for notified MDR/RR-TB. Although the primary ambition was to assess whether evidence of transmission could be approached from readily observable data—which we expect to be more relevant for policymakers—we assessed the model's sensitivity to potential underreporting that may be seen in notified cases. Moreover, we assessed the impact of potentially underreported treatment, by multiplying the number of MDR/RR-TB receiving BDQ by the ratio of estimated to notified MDR/RR-TB.

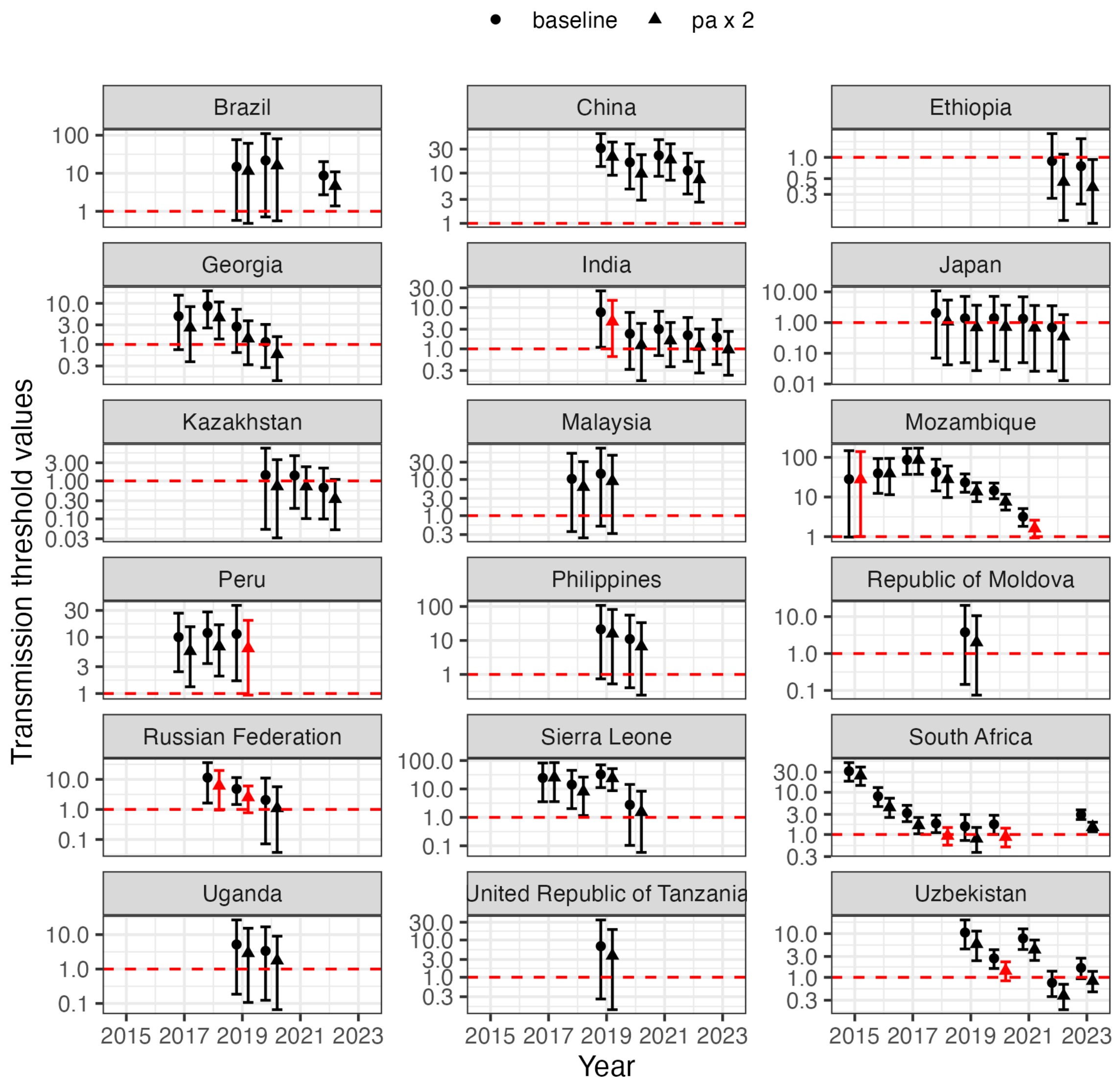


**Figure S3:** sensitivity analysis varying the probability of acquired resistance. Black circles are the baseline scenario. Triangles are acquired resistance increased by a factor of 2 from baseline values. Highlighted in red where sensitivity analysis changed the qualitative interpretation (lower bound of 95% credible interval above/below 1). Points are median estimates. Error bars are 95% credible intervals. Note, y-axes differ between subplots and shown on a logarithmic scale.

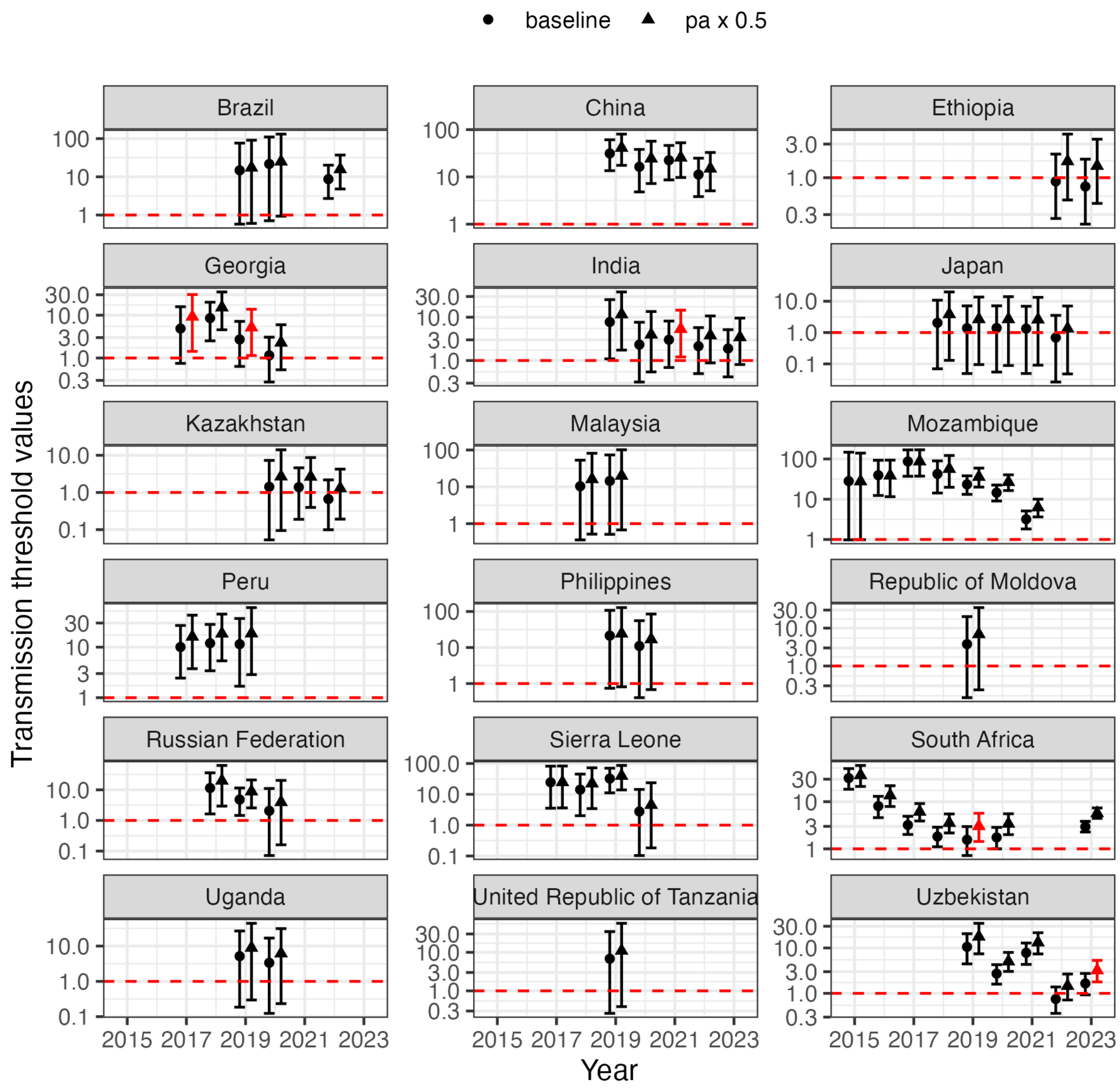


**Figure S4:** sensitivity analysis varying the probability of acquired resistance. Black circles are the baseline scenario. Triangles are acquired resistance decreased by a factor of 2 from baseline values. Highlighted in red where sensitivity analysis changed the qualitative interpretation (lower bound of 95% credible interval above/below 1). Points are median estimates. Error bars are 95% credible intervals. Note, y-axes differ between subplots and shown on a logarithmic scale.

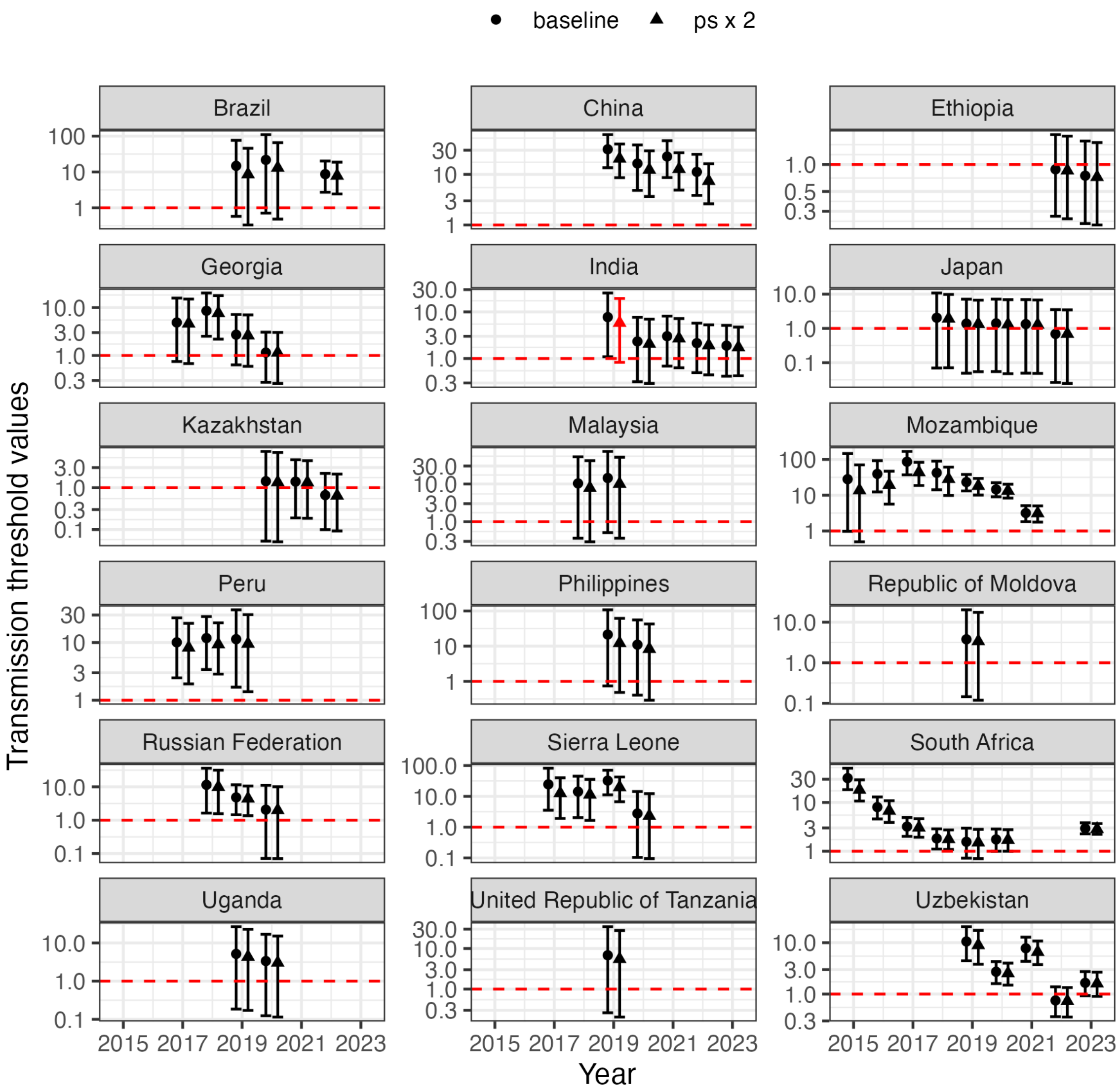


**Figure S5:** sensitivity analysis varying the probability of spontaneous resistance. Black circles are the baseline scenario. Triangles are spontaneous resistance increased by a factor of 2 from baseline values. Highlighted in red where sensitivity analysis changed the qualitative interpretation (lower bound of 95% credible interval above/below 1). Points are median estimates. Error bars are 95% credible intervals. Note, y-axes differ between subplots and shown on a logarithmic scale.

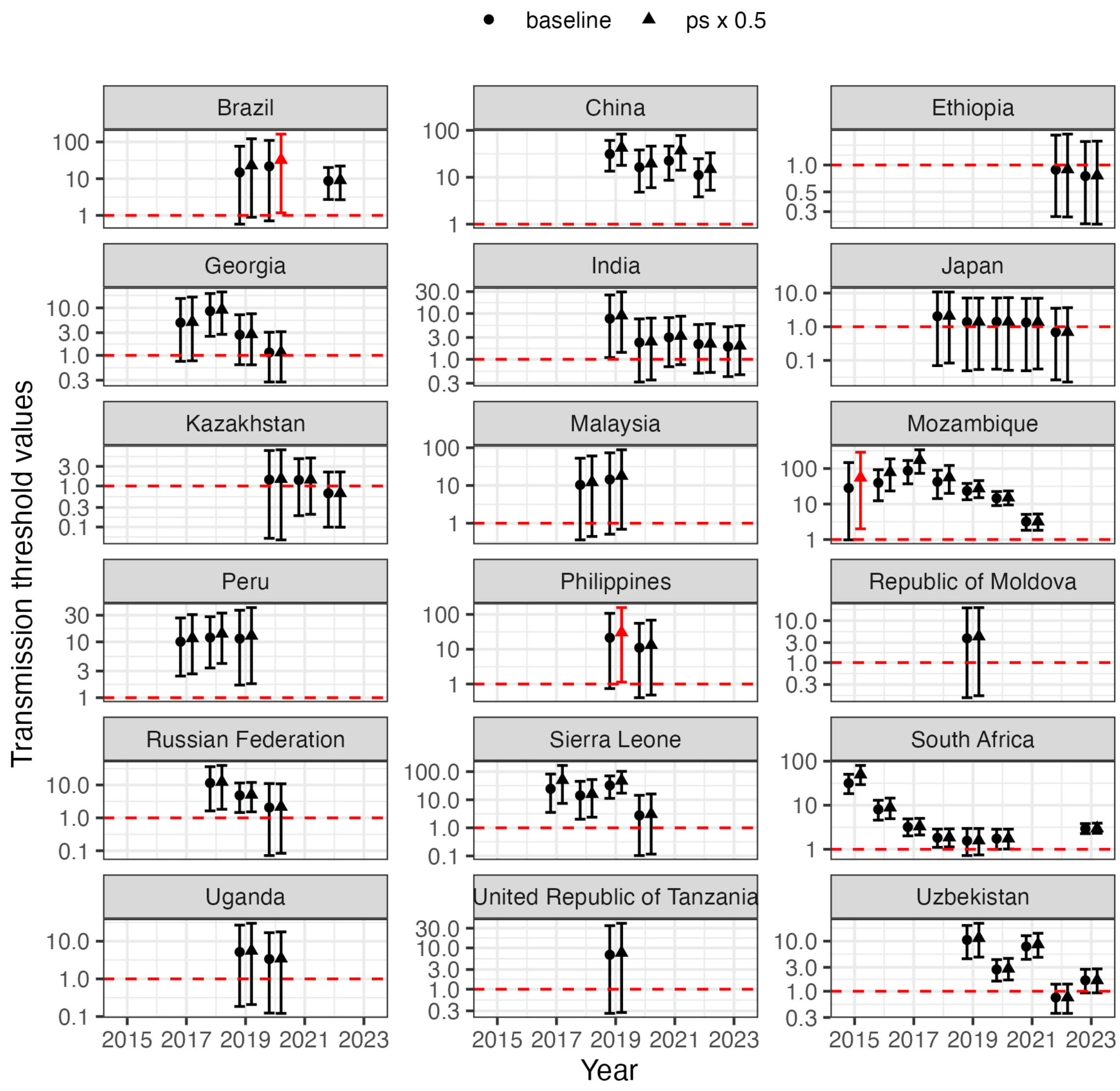


**Figure S6:** sensitivity analysis varying the probability of spontaneous resistance. Black circles are the baseline scenario. Triangles are spontaneous resistance decreased by a factor of 2 from baseline values. Highlighted in red where sensitivity analysis changed the qualitative interpretation (lower bound of 95% credible interval above/below 1). Points are median estimates. Error bars are 95% credible intervals. Note, y-axes differ between subplots and shown on a logarithmic scale.

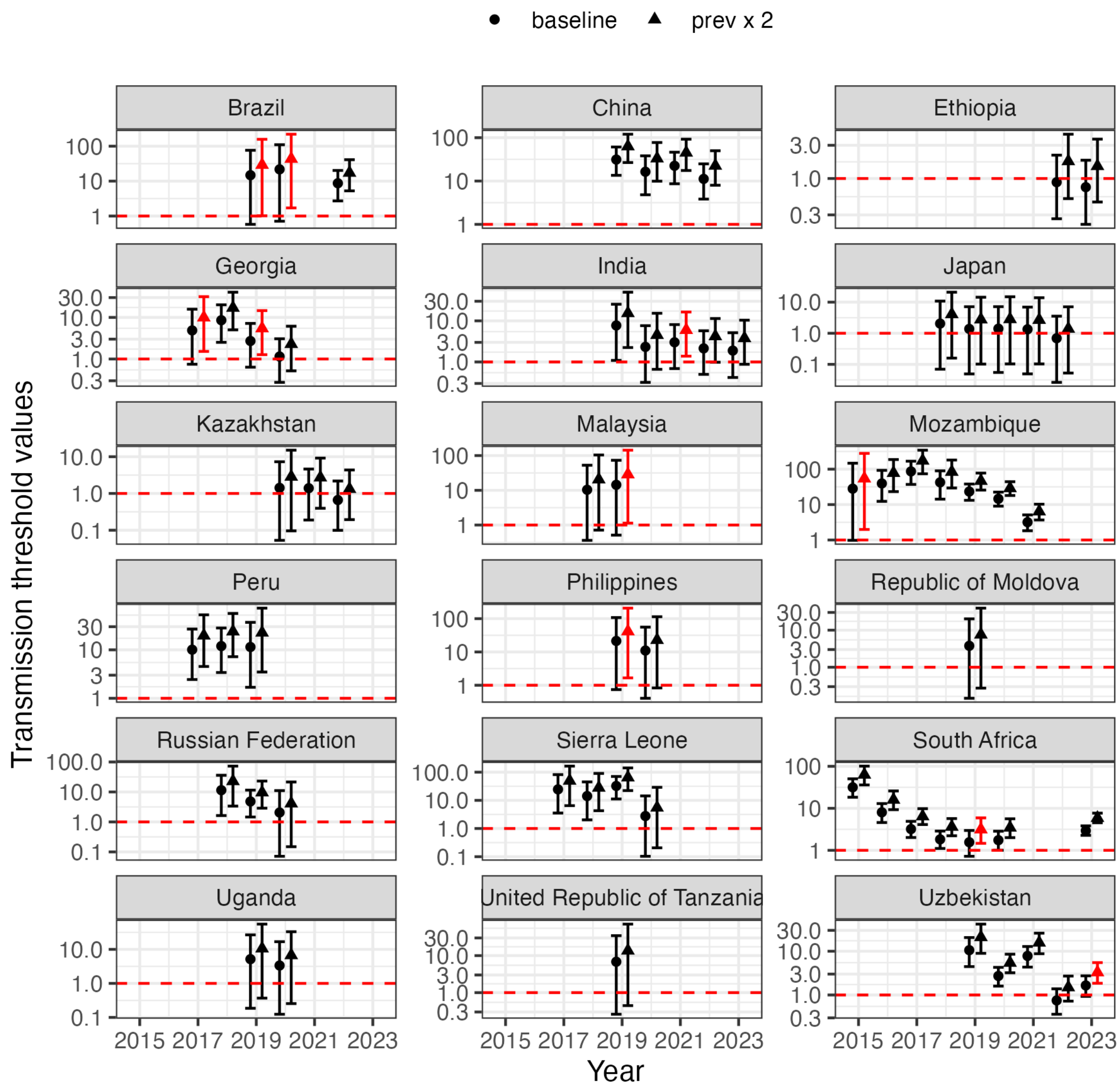


**Figure S7:** sensitivity analysis varying the prevalence of BDQR among MDR/RR-TB. Black circles are the baseline scenario. Triangles are prevalence increased by a factor of 2 from baseline values. Highlighted in red where sensitivity analysis changed the qualitative interpretation (lower bound of 95% credible interval above/below 1). Points are median estimates. Error bars are 95% credible intervals. Note, y-axes differ between subplots and shown on a logarithmic scale.

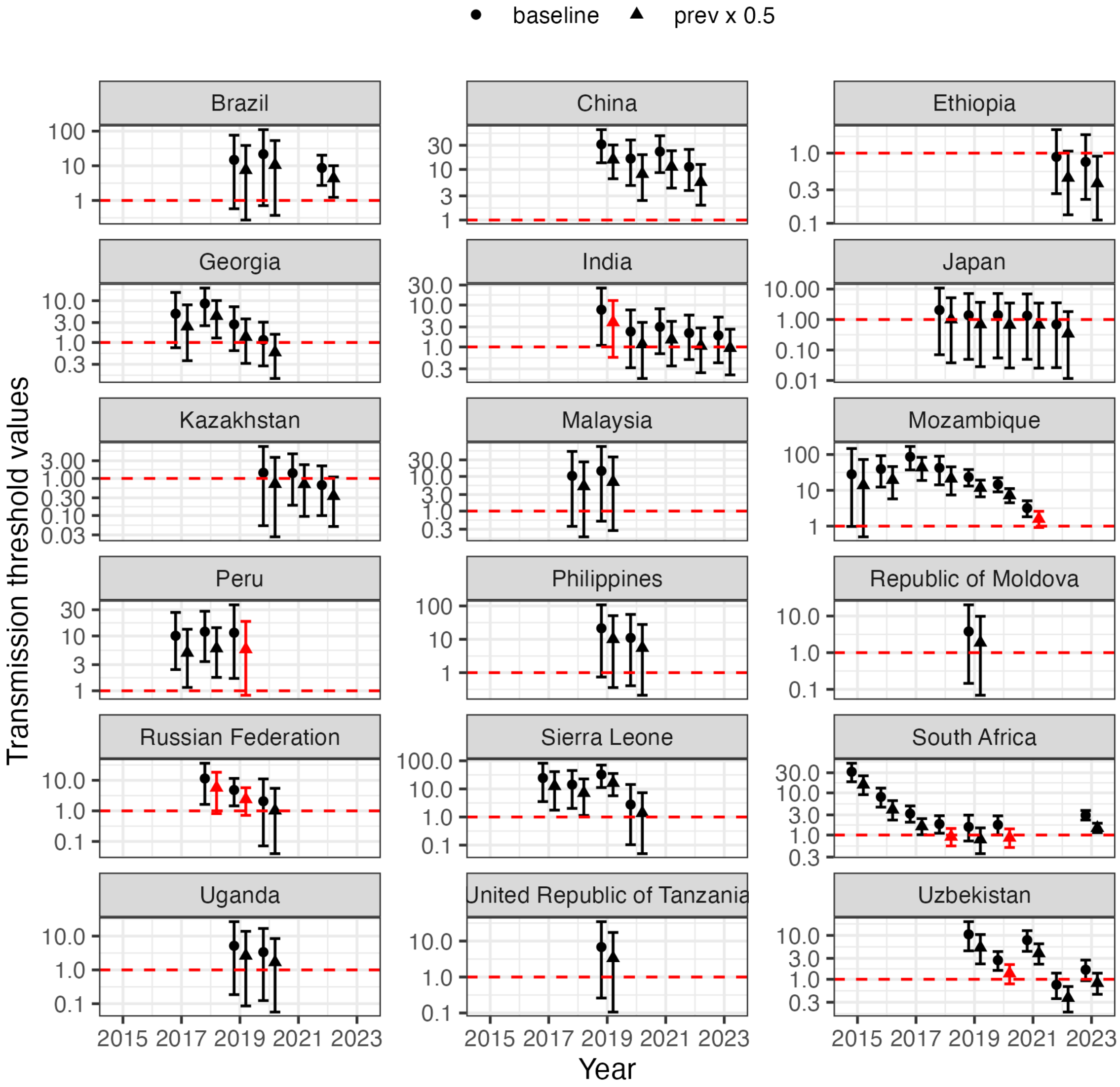


**Figure S8:** sensitivity analysis varying the prevalence of BDQR among MDR/RR-TB. Black circles are the baseline scenario. Triangles are prevalence decreased by a factor of 2 from baseline values. Highlighted in red where sensitivity analysis changed the qualitative interpretation (lower bound of 95% credible interval above/below 1). Points are median estimates. Error bars are 95% credible intervals. Note, y-axes differ between subplots and shown on a logarithmic scale.

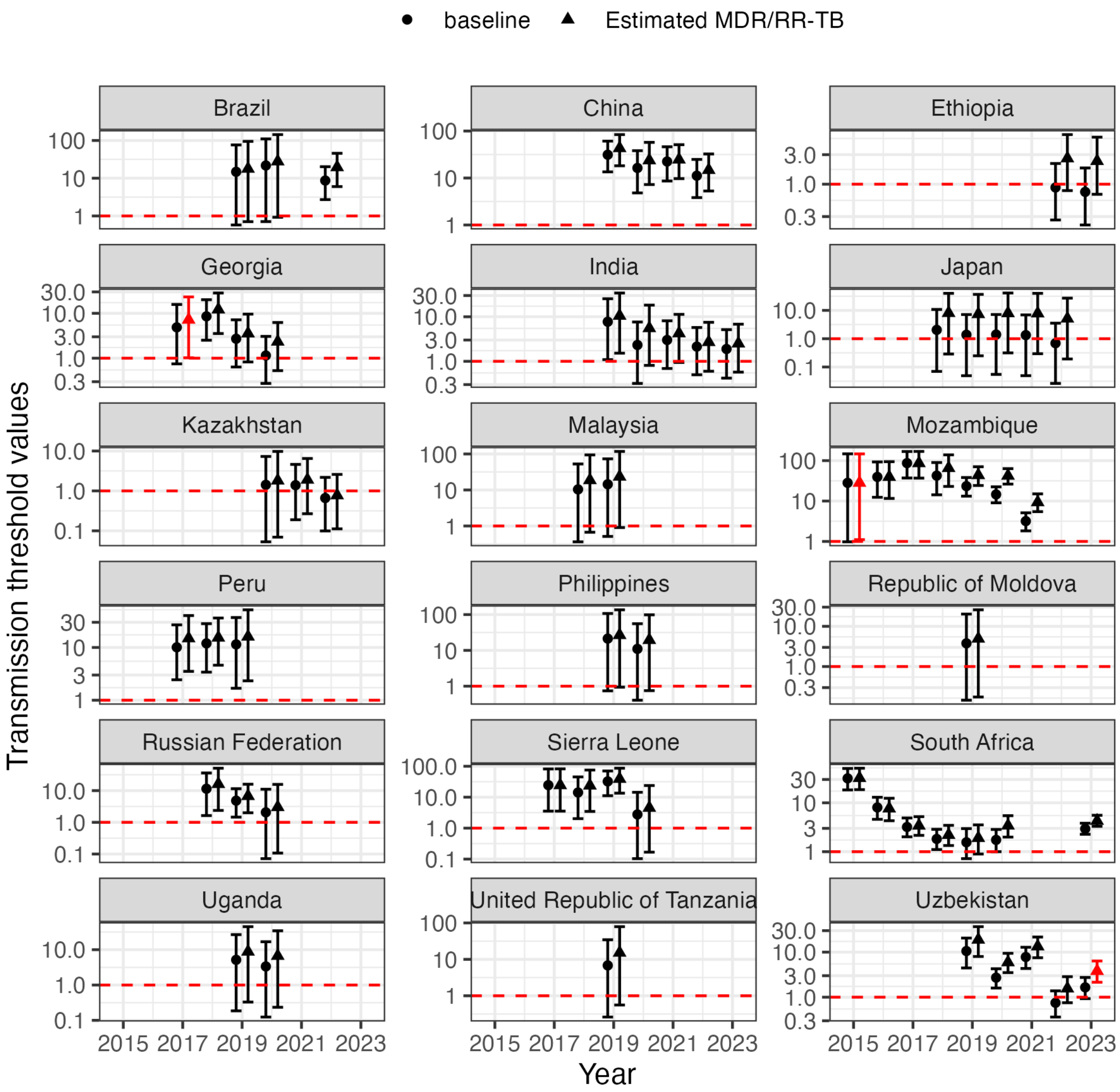


**Figure S9:** sensitivity analysis of varying MDR/RR-TB input data. Black circles are WHO notified MDR/RR-TB. Triangles are WHO estimated MDR/RR-TB. Highlighted in red where sensitivity analysis changed the qualitative interpretation (lower bound of 95% credible interval above/below 1). Points are median estimates. Error bars are 95% credible intervals. Note, y-axes differ between subplots and shown on a logarithmic scale.

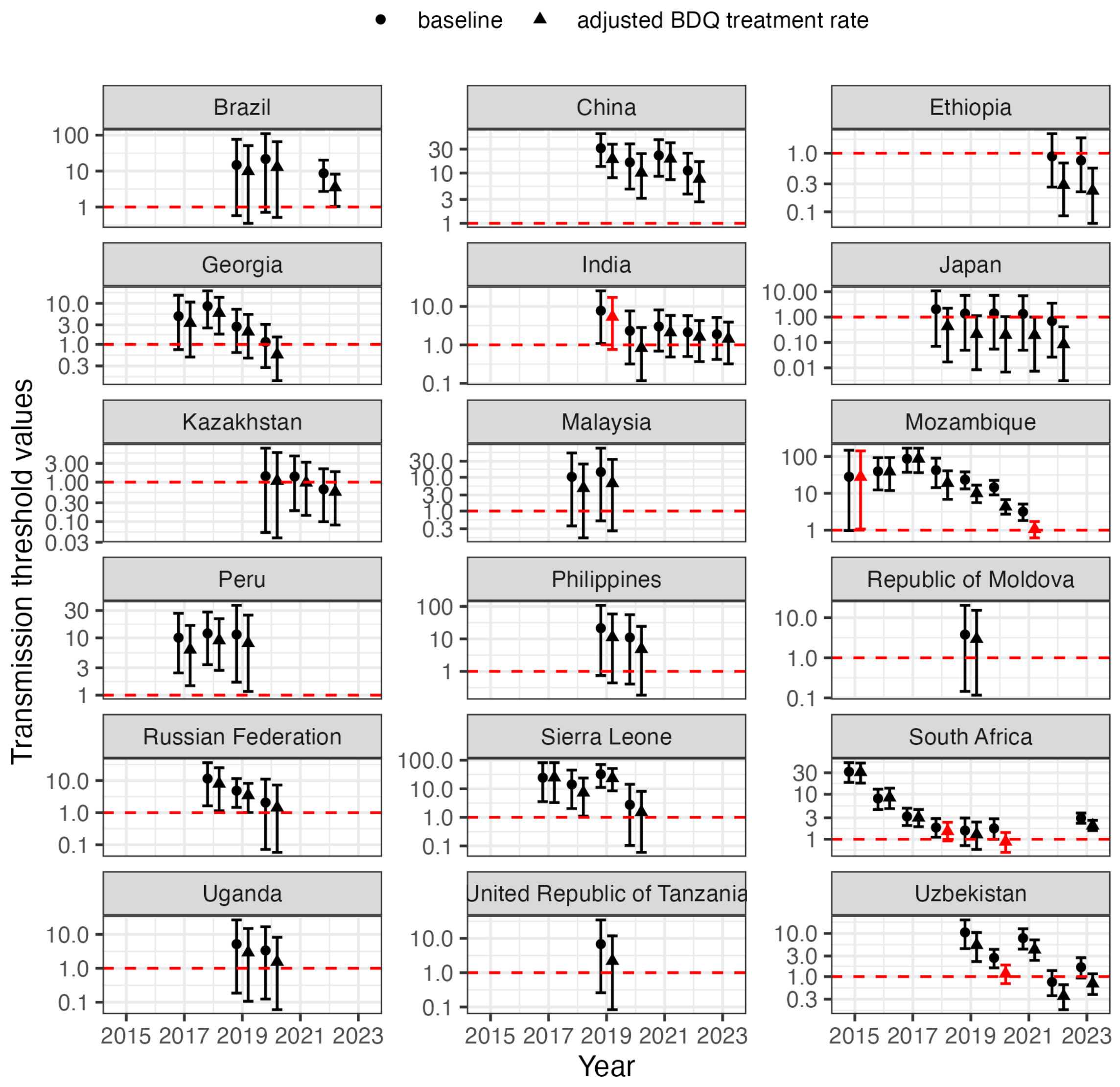


**Figure S10:** sensitivity analysis of varying BDQ treatment input data. Black circles are WHO notified BDQ treatment rate. Triangles are adjusted BDQ treatment rate. Highlighted in red where sensitivity analysis changed the qualitative interpretation (lower bound of 95% credible interval above/below 1). Points are median estimates. Error bars are 95% credible intervals. Note, y-axes differ between subplots and shown on a logarithmic scale.

### References

1. Kendall EA, Fofana MO, Dowdy DW. Burden of transmitted multidrug resistance in epidemics of tuberculosis: a transmission modelling analysis. Lancet Respir Med. 2015;3: 963–972. doi:10.1016/S2213-2600(15)00458-0

2. Nguyen TVA, Anthony RM, Bañuls A-L, Nguyen TVA, Vu DH, Alffenaar J-WC. Bedaquiline Resistance: Its Emergence, Mechanism, and Prevention. Clin Infect Dis. 2018;66: 1625–1630. doi:10.1093/cid/cix992

3. Degiacomi G, Sammartino JC, Sinigiani V, Marra P, Urbani A, Pasca MR. In vitro Study of Bedaquiline Resistance in Mycobacterium tuberculosis Multi-Drug Resistant Clinical Isolates. Front Microbiol. 2020;11: 559469. doi:10.3389/fmicb.2020.559469

4. Andries K, Villellas C, Coeck N, Thys K, Gevers T, Vranckx L, et al. Acquired Resistance of Mycobacterium tuberculosis to Bedaquiline. Van Veen HW, editor. PLoS ONE. 2014;9: e102135. doi:10.1371/journal.pone.0102135

5. Ismail N, Sirgel F, Omar SV, Omar S, De Kock M, Spies C, et al. Unpacking bedaquiline heteroresistance: the importance of intermediate profiles for phenotypic drug susceptibility testing. Shields RK, editor. Antimicrob Agents Chemother. 2025; e00356-25. doi:10.1128/aac.00356-25

6. Colijn C, Cohen T, Ganesh A, Murray M. Spontaneous Emergence of Multiple Drug Resistance in Tuberculosis before and during Therapy. Mokrousov I, editor. PLoS ONE. 2011;6: e18327. doi:10.1371/journal.pone.0018327

7. Hu X, Wu Z, Lei J, Zhu Y, Gao J. Prevalence of bedaquiline resistance in patients with drug-resistant tuberculosis: a systematic review and meta-analysis. BMC Infect Dis. 2025;25: 689. doi:10.1186/s12879-025-11067-2

8. Mallick JS, Nair P, Abbew ET, Van Deun A, Decroo T. Acquired bedaquiline resistance during the treatment of drug-resistant tuberculosis: a systematic review. JAC-Antimicrob Resist. 2022;4: dlac029. doi:10.1093/jacamr/dlac029
